# The Safety of Intravenous Lipopolysaccharide for the Study of Systemic Inflammation in Humans: A Scoping Review

**DOI:** 10.1101/2025.06.16.25329686

**Authors:** Unimark Junior S. Awadzi, Ryan A. Heumann, Grzegorz B. Gmyrek, J. Paul Robinson, Matthew P. Ward, Steven R. Steinhubl

## Abstract

**Background and Objective:** This review was conducted to evaluate the safety of intravenous lipopolysaccharide (IV LPS) for human immunology research through a scoping assessment of published clinical trials and pertinent case reports. As a collection of case studies reporting rare instances of sinus pauses following IV LPS had been reported in 2005, along with suggested approaches to minimize them, we limited our review to articles published since 2005 in order to focus on contemporary safety.

**Method:** PRISMA Extension for Scoping Reviews (PRISMA-ScR) guidelines were followed. English language articles meeting inclusion criteria were identified in PubMed through February 2025.

**Results:** From 234 articles we identified 155 distinct studies, with the majority (n=113) limiting enrollment to males only. These studies included a total of 3,551 volunteers, almost exclusively healthy volunteers, with an estimated mean age of 26.0 (range 20 to 56). In all studies, IV LPS elicited a dose-dependent inflammatory response characterized by elevations of heart rate, body temperature, and influenza-like symptoms that typically resolved by 8 hours post dosing. The only unanticipated serious adverse events identified were 2 cases of mild syncope associated with 4-5 second sinus pauses that both occurred in a single study.

**Conclusion:** The rarity of adverse events across the 155 studies reviewed suggests that the LPS model of systemic inflammation in humans is safe and well tolerated, although precautions should be taken to minimize vagally induced sinus pauses.

## I. Introduction

There are few systems in the human body more complex or more individualized than the immune system [1]. Exposure to the same exact pathogen in two different people can lead to different immune responses due to this inter-individual variability [2]. Genetic variation, age, previous pathogen exposures, sex, nutrition, environmental exposure, and the human microbiome can all influence individual immune response [3] [4] [5] [6]. This complexity, and the critical role our immune systems plays in a range of life-threatening health conditions including infections, cancer, autoimmune diseases, and more, underscores the importance of improving our understanding of individual differences in immune response.

Intravenous lipopolysaccharide (IV LPS), also referred to as endotoxin, is a well-established model for the controlled study of innate immune system activation and subsequent inflammation in humans. For many decades now, the model has been used in hundreds of studies related to human immune and inflammation responses [7] [8] [9] [10], endocrine and metabolic systems [11] [12] [13] [14], psychological and neural processes [15] [16] [17] [18], microbiota and gut health [19] [20] [21], and the study of various cellular and molecular mechanisms [22] [23] [24] [25]. While the intravenous LPS model has been most commonly used [26], other routes like intradermal [27], and intrabronchial [28] are also increasingly employed in the study of organ specific inflammation, but these alternative delivery routes will not be covered in this review.

In LPS studies carried out prior to 2006, a total of 5 cases of temporary sinoatrial node conduction abnormalities manifesting as asystole, sinus pauses, or severe bradycardia have been reported [29] [30] [31]. The first case involved a 28-year-old volunteer who experienced a sinus pause of approximately 23 seconds with a temporary loss of consciousness approximately an hour after receiving a 2 ng/kg E. coli 0113 IV bolus of LPS.[29] Fluid administration with magnesium sulfate was used as an intervention. In retrospect it was determined that the volunteer had a history of severe vasovagal syncope, and he had fasted prior to the IV LPS and was likely dehydrated. Another report, including a total of 4 cases (3 new cases plus the previously reported case [29]), was then published describing events in volunteers following 2 - 4 ng/kg of LPS (E. coli 0113), with sinus pauses ranging from 4 up to the previously reported 23 seconds. [30] In these cases, interventions included brief chest compression, atropine and fluid challenges. It was found that three of the 4 cases had a prior history of syncope. One additional case was reported, out of over 650 volunteers who received 2 - 4 ng/kg of LPS (E. coli 0113), in which a sinus pause lasting several seconds occurred during an episode of nausea and vomiting. [31] Based on these cases, it was recommended that volunteers for IV LPS studies going forward should be screened for a history of syncope and should be hydrated before and during the LPS challenge.

While several excellent general reviews of the LPS model of human inflammation have been published over the last few decades, a comprehensive review of safety outcomes has not previously been published [32] [33] [34] [35] [36]. A contemporary safety review is especially important following the publication of the collection of case reports of cardiac conduction abnormalities in 2005 and recommendations as to how to minimize them. Therefore, we conducted a search of all English language articles describing clinical studies using IV LPS in humans since 2005. Our objective was to identify all reported adverse reactions and summarize all potential risks or safety issues associated with contemporary human IV LPS studies.

## II. Method

PRISMA Extension for Scoping Reviews (PRISMA-ScR) guidelines were followed and Covidence.org was used as an auxiliary tool. Between November 2023 and February 2025, 3,156 published manuscripts indexed in PubMed of potential interest were identified using Medical Subject Headings (MeSH) terms for “Endotoxins”, “Endotoxemia”, “Lipopolysaccharide”, “LPS”, “Intravenous”, “Injections”, “IV”, “Infusions”, “Humans”, and “Bolus.” The Covidence website (https://www.covidence.org/) provided the platform for screening the articles for the determination of appropriateness for inclusion for review. The initial criteria required the article to be published in English and include intravenous administration of LPS to humans. On the Covidence website, two screening steps ensued after references were imported. (**Figure 1**) First, the title and abstracts of the 3,156 initially identified papers were screened to determine the articles that demonstrated the use of LPS in humans, which led to the discovery of 506 articles that matched our search terms. These articles were downloaded from their sites of publication and uploaded to the website as PDFs and then underwent manual full text screening. After full text screening, a final set of 234 articles, published after 2005, were included in this scoping review. Each underwent manual review, with data extraction carried out and compiled into a table that included essential elements such as study designs, participant characteristics, and safety outcomes. (**Supplementary Table**) As several studies generated multiple publications, efforts were made to ensure that volunteers were not recounted, but that all unique findings from each study were captured. As a result, 155 primary IV LPS studies were identified from the 234 articles.

**Fig 1.**
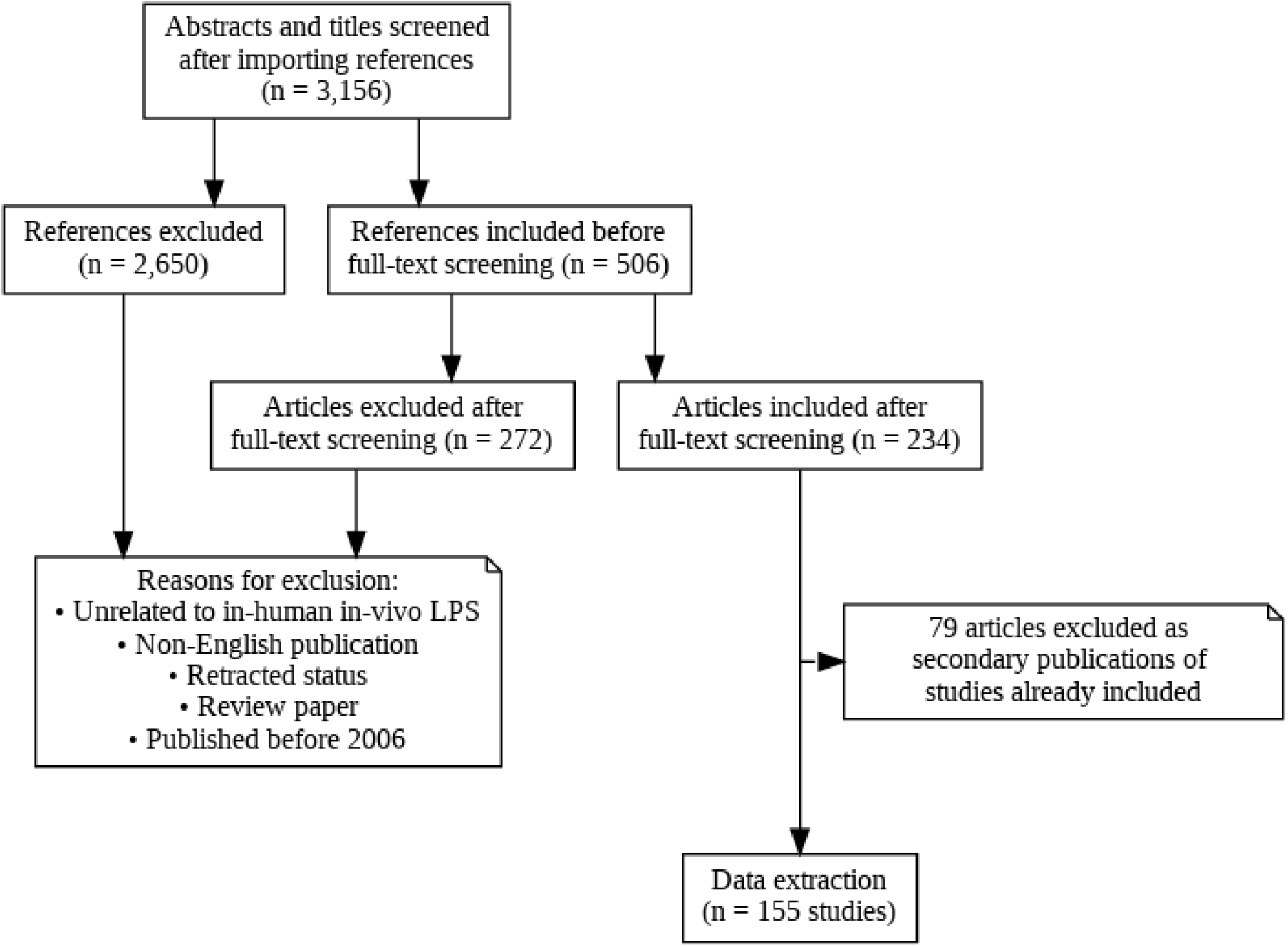
Flow chart of the screening processes to include IV LPS studies published between 2006 and February 2025.

## III. Results

### Volunteer characteristics

Between 2006 and February 2025, a total of 3,551 volunteers received IV LPS across 155 published studies. Per study, the median number of participants who received IV LPS was 16.0, with a mode of 8 (8.4%). The largest study included 294 volunteers [37]. The studies almost exclusively recruited healthy volunteers (n =154 studies, 99.4%), with 113 studies (73%) including only male volunteers. Two studies included only females and the remaining recruited both males and females (n = 40 studies). The overall mean (+standard deviation) age of volunteers was 26.0 ± 0.98 years with the vast majority of volunteers under 37 years age. (**Figure 2**).

**Fig 2.**
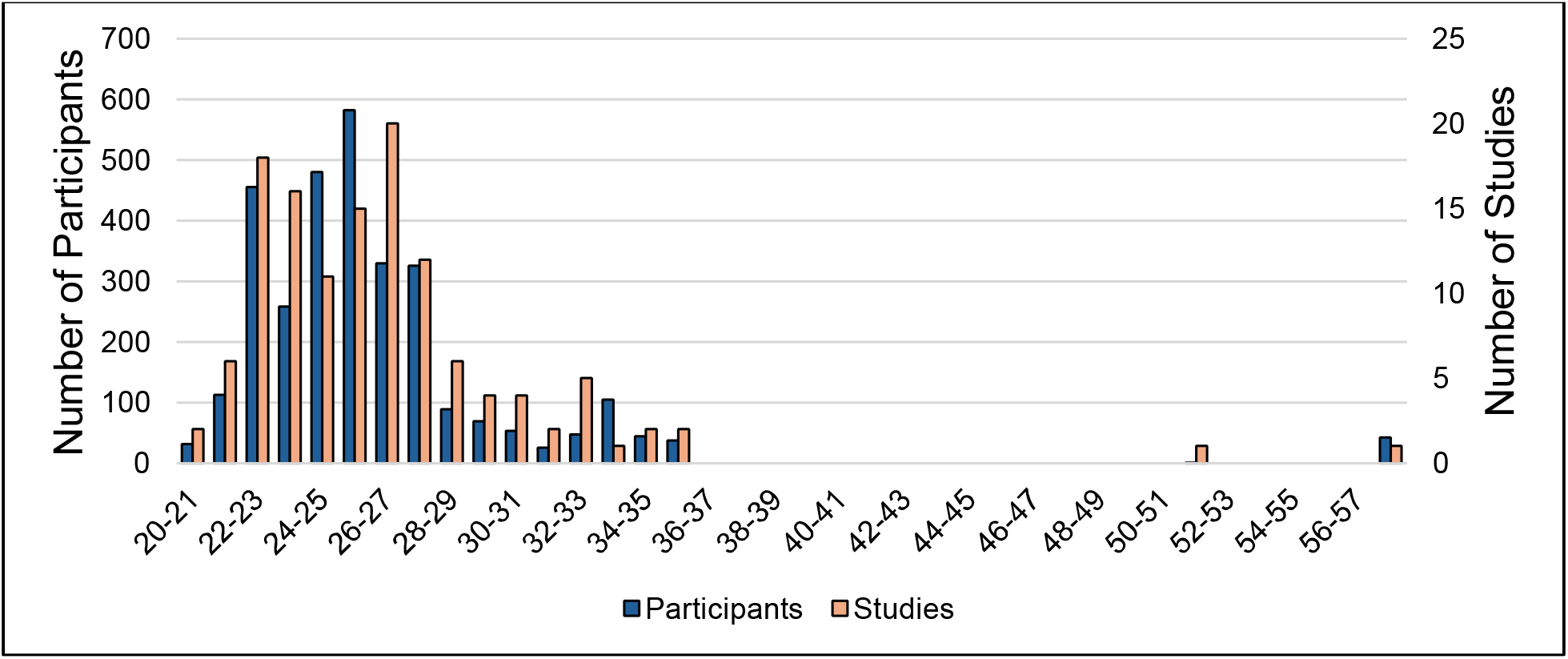
Estimated mean age distribution of all studies. The overall mean age is 26.0 (see supplementary methods for calculation). The right y-axis represents the number of studies with a reported mean age, or surrogate mean age, between the age ranges on the x-axis. The left y-axis represents the sum of the number of participants in all those studies.

### Relationship Between LPS Dose on Biomarkers of Inflammation and Symptoms of Sickness

The lowest IV bolus dose recorded among the included studies was 0.1 ng/kg, the highest dose was 4 ng/kg, and the mode was 2 ng/kg. Several studies have evaluated the dose response of LPS. In one study aimed at identifying the minimal dose of endotoxin needed to initiate observable changes in various markers of inflammation, saline (placebo) or LPS doses of either 0.1, 0.25, 0.5, 1.0, or 2.0 ng/kg were administered to healthy human volunteers with ~4 volunteers per dose [38]. Neither placebo nor the 0.1 ng/kg cohort showed any significant change from baseline for temperature, heart rate, white blood cell (WBC) count, serum IL-6, or serum TNF-α (**Table 1**). Meanwhile, the 1.0 ng/kg and 2.0 ng/kg cohorts led to significantly higher and faster temperature peaks compared to 0.25 ng/kg and 0.5 ng/kg and were the only doses to lead to reactions meeting systemic inflammatory response syndrome (SIRS) criteria for temperature (over 38°C) and heart rate (over 90 bpm).(**Table 1**) No significant change or pattern in mean arterial pressure was seen for any dose.

**Table 1.** Maximal increases in temperature, heart rate (HR), serum IL-6, and serum TNF-α following different doses of IV LPS compared to baseline. From Herlitz et al. (2015) [38].

| Study Group | Physiological parameters and cytokines measured |  |  |  |
| --- | --- | --- | --- | --- |
| | Temp. ( °C) | HR (bpm) | IL-6 (pg/mL) | TNF-α (pg/mL) |
| Placebo | - | - | - | - |
| 0.1 ng/kg | - | - | - | - |
| 0.25 ng/kg | 0.7 ± 0.4 (p<0.02) | - | - | - |
| 0.5 ng/kg | 0.8 ± 0.3 (p<0.02) | 23 ± 7 (p<0.02) | 82 (p<0.02) | 53 ± 18 (p<0.01) |
| 1.0 ng/kg | 1.6 ± 0.1 (p<0.02) | 42 ± 1 (p<0.05) | > 300* (p<0.02) | 215 ± 29 (p<0.01) |
| 2.0 ng/kg | 1.6 ± 0.2 (p<0.02) | 35 ± 5 (p<0.05) | >300* (p<0.02) | 290 ± 56 (p<0.01) |

“- “represents insignificant change from baseline.
\* - peak detectable level of IL-6 was 300 pg/ml
p-values indicate the statistical significance levels of these results relative to baseline.

Not surprisingly, the frequency and severity of symptoms experienced also appears to be dose dependent. While no contemporary studies have evaluated the relationship between symptoms and dose, a 1999 study of 20 individuals receiving 0-, 1-, 2-, and 4-ng/kg doses of intravenous E. coli O113 LPS, found that adverse experiences (e.g., chills, myalgia, headache, nausea, or vomiting) were most common with the highest dose, with vomiting occurring in 2 people – one receiving the 2 ng/kg dose and the other a 4 ng/kg dose [39]. Flu-like symptoms reported in studies typically start approximately 1 hour after the LPS challenge and usually resolve within 8 hours. In studies that tracked flu-like responses, no common IV LPS dose has been shown to not illicit these symptoms.

### Unanticipated Adverse Events

Adverse events beyond flu-like symptoms were rarely reported after IV LPS experiments, with only 4 reported episodes out of 3, 551 IV LPS volunteers in 155 studies. Of these, the only reported unanticipated adverse event that investigators attributed to LPS was 2 cases of temporary cardiac sinoatrial node dysfunction (discussed below). Other reported adverse events but concluded by investigators to not be due to LPS, included one case of delayed onset of Type 1 diabetes, but retrospective analysis of baseline blood samples identified high levels of diabetes-specific autoantibodies in that individual prior to their receiving LPS [40]. One other study reported a case of acute gastroenteritis, but the investigators attributed this to familial transmission [41] [42].

#### a. Sinoatrial Node Dysfunction

Among the 155 studies, the only significant adverse event attributed to LPS was brief syncope associated with 4-5 second sinus pauses in two volunteers, both occurring in a single study [43].

As noted earlier, this has been previously reported, but these are the only reported cases in the 20 years since those initial reports. Although these cases occurred in the same study, there was no clear participant-specific or methodologic explanation.

To fully explore all that is known of sinus node dysfunction secondary to a presumed vagal response to IV LPS, we summarized all cases ever reported in the English literature in **Table 2**. Including the 2 cases identified in this review, there has been a total of 7 cases in the literature, with the first reported in 2000 [29]. All reported cases were short-lived with no long term sequalae.

Based on these 7 reported cases, in an estimated 6, 441 IV LPS-exposed individuals (3, 551 from the current analysis and an estimated 2,890 from pre-2006 reports), we would estimate the incidence of temporary sinus node dysfunction in IV LPS to be 0.11%, or ~1 per 1000 LPS-treated volunteers. Our finding of only 2 cases among 3,551 LPS-challenged volunteers since assumed screening and hydration practices suggests that with contemporary methods that the incidence is even less, ~1 per 1,800 volunteers.

**Table 1.**
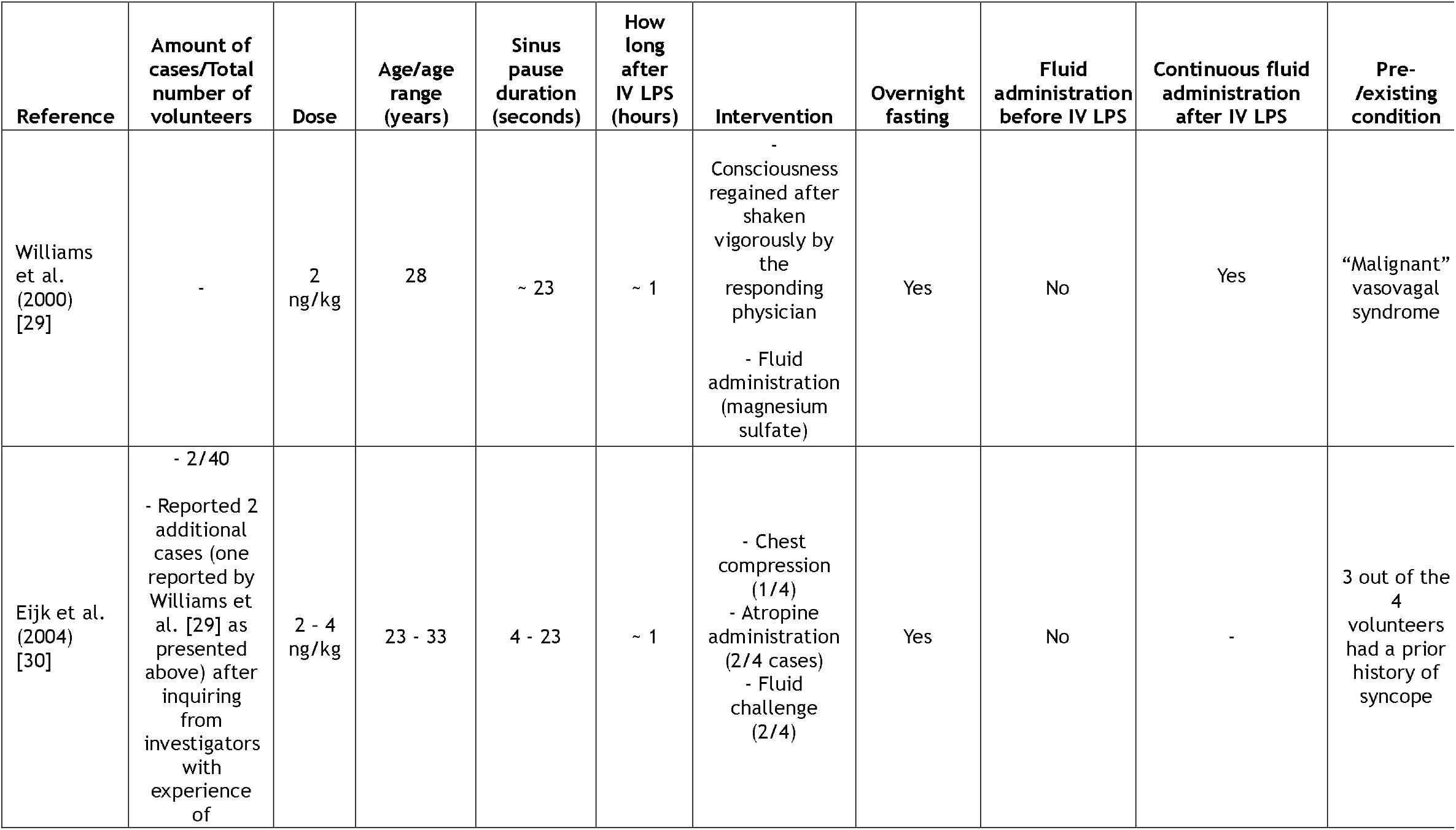
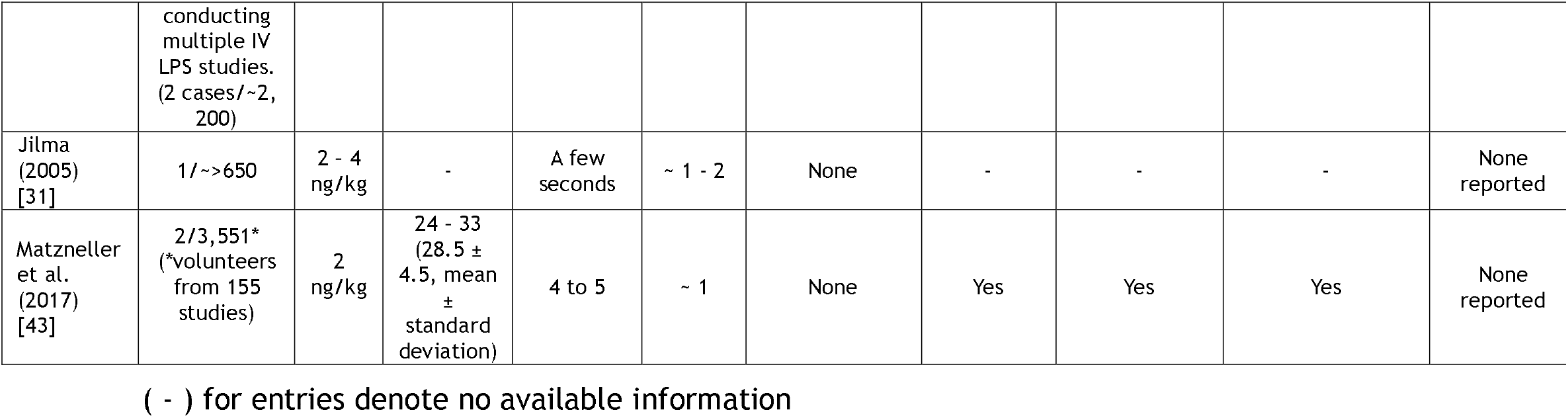
Summary of all cardiac conduction abnormally cases.

## IV. Discussion

To minimize the inadequacies of animal models, human models of systemic inflammation are essential for accurately identifying disease-related mechanisms. While animal models have been extensively used to study immune activation and inflammation, there are limitations to their translation to humans [44] [45] [46] [47] [48] [49]. There are notable differences in the immune responses to LPS challenges between humans and animals, as some facets of human immunology cannot be accurately replicated in other species [49] [50]. Species-specific sensitivity to LPS varies, with humans being the most sensitive among all species [51] [52]. The dose of LPS used in most *in vivo* studies on mice typically ranges from 1 to 25 mg/kg, a dosage that is one-million times the 2 – 4 ng/kg dose of E. coli O113-derived LPS commonly utilized in humans [53]. Beyond sensitivity, the immune response to LPS in mice also differs from humans since some chemokines are unique to humans and not found in mice, while others are present in mice but not in humans [50]. Also, mouse models’ genomic response correlates poorly with that in humans [44] [49].

This is, to the best of our knowledge, the largest comprehensive review of the safety of IV LPS in human volunteers. Beyond the expected flu-like symptoms, we found IV LPS to be safe. The only reported unanticipated adverse event being rare (~1 in 1000 overall, and ~1 in 1800 exposed individuals in contemporary practice) cases of temporary sinus node dysfunction, presumed to be vagally mediated, and manifesting as sinus pauses lasting ~ 4 – 23 seconds. This complication, while rare, should be considered as a potential, but addressable risk of IV LPS in volunteers. To minimize the risk of temporary sinus node dysfunction, excluding individuals with a history of vasovagal syncope or near syncope, minimizing nausea and vomiting by using the lowest LPS doses necessary, and assuring adequate hydration are important.

### Limitations

We are only able to make safety conclusions regarding LPS in a generally younger population as the mean age of volunteers included in our analysis was only 26. A 2001 study compared the response to 2 ng/kg E. coli LPS in 9 individuals aged 61 – 69 years compared to 8 individuals aged 20 – 27 [54]. They found that the older cohort experienced greater inflammation based on a prolonged fever response and a larger increase in some serum inflammatory biomarkers. This could suggest that elderly volunteers might be at higher risk for experiencing greater symptoms. There is one contemporary study that recruited participants aged 55 to 90 years receiving a 0.4 ng/kg LPS bolus but was reported only as an abstract with no report of safety events, and therefore not included in this review [55].

The reported number of volunteers evaluated, 3,551, does not accurately reflect the total number of individuals involved in IV LPS studies since the model’s inception, as we limited our analysis only to studies published after 2005. Additionally, as we limited our review to English-language, peer-reviewed published articles, there are likely even contemporary LPS-treated individuals who were not included in this analysis. Also, although we made every effort to ensure that volunteers were not recounted due to multiple publications from the same study, it is possible some duplication might have occurred.

## V. Conclusion

In this review, encompassing all published English-language IV LPS studies from the last 20 years, we found the model to be safe and well tolerated. However, vagally mediated temporary sinus node dysfunction can be a rare, self-limited, serious adverse reaction to IV LPS. The risk can be minimized through careful participant selection, assuring adequate hydration throughout, and minimizing nausea.

## Data Availability

All data produced in the present work are contained in the supplementary documents

## Acknowledgement

Special thanks to Professor Jane Yatcilla from Purdue University Science and Engineering Libraries Administration for providing the relevant resources for the literature search on the PubMed database.

## Supplemental Methods

The overall mean age, or the estimated mean age of all participants of all studies, was calculated using the reported means or medians of each study individually. Seven studies did not include any information regarding age, and 18 only mentioned age ranges, so all 25 were excluded from the calculation. Mean was used unless only the median was reported, in which case it was used as a surrogate. For calculation, the mean, or surrogate mean, of each study was multiplied by the number of participants of that study. These products were summed for all studies and divided by the total number of participants to give the overall mean age. Population standard deviation was then calculated by summing the squared differences of each study mean age and the overall mean age, dividing the result by the total number of participants, and taking the square root.

## Declarations

### Clinical trial number

not applicable.

Human Ethics and Consent to Participate declarations: not applicable.

### Consent to Participate declaration

not applicable.

### Approval Committee or the Internal Review Board (IRB) in the Ethics Approval declaration

not applicable.

### Clinical trial number

not applicable.

