## Supplementary Table for "The Safety of Intravenous Lipopolysaccharide for the Study of Systemic Inflammation in Humans: A Scoping Review"

| **STUDY NAME (PUBLICATION AND REFERENCE)** | **PRIMARY RESEARCH OBJECTIVES** | **NUMBER OF PARTICIPANTS** | **PARTICIPANT CHARACTERISTICS** | **AGE RANGE (Years)** | **DOSES USED** | **MEASURED RESPONSES TO LPS** | | | | **TOTAL DURATION OF MONITORING** | **SAFETY CONCERNS NOTED - ANY**  **DROPOUTS?** | **LOCATION** |
| --- | --- | --- | --- | --- | --- | --- | --- | --- | --- | --- | --- | --- |
|  |  |  |  |  |  | **PHYSIOLOGICAL PARAMETERS MONITORED** | **CYTOKINES MEASURED** | **SUBJECTIVE SYMPTOMS RECORDED** | **KEY FINDING** |  |  |  |
| [1] Physiologic variability at the verge of systemic inflammation:  multi-scale entropy of heart rate variability is affected by very  low doses of endotoxin  [unique] | Assessed Multi-scale Entropy’s effectiveness in detecting inflammation after bolus injection.  The study also aimed at identifying the minimal doses of endotoxin needed to initiate observable changes in various inflammation markers and juxtapose HRV metrics sensitivity to the induced changes | 25  Females (n = 8)  Males (n = 17)  LPS = 21 | Healthy human volunteers (Male, Female) | 19 – 31  Mean age = 25 | 0.1, 0.25, 0.5, 1.0, or 2.0 ng/kg  E. coli  Clinical Center Reference Endotoxin  NIH  Bethesda, Md  CC-RE, lot 2 | Heart rate, temperature, mean arterial blood pressure (MAP) | TNF-α, and IL-6 | None | Increased temperature readings (dose-dependent)  Increased heart rate (dose-dependent)  White blood cell count rose (dose-dependent)  Interleukin 6 increased (among the 1 and 2 ng/kg groups)  TNF-α increased (within the 0.5, 1 and 2 ng/kg groups)  The 2 ng/kg group experienced a decrease in SDNN (Standard Deviation of NN intervals).  The 1 or 2 ng/kg groups experienced a decrease in PNN50  The 1 or 2 ng/kg groups experienced a decrease in RMSSD (Root Mean Square of Successive Differences)  The 2 ng/kg group experienced a rise in low frequency/high-frequency (LF: HF) ratio  The 1 and 2 ng/kg had a decrease in MSE (Multiscale entropy) curves which depends on the dose of LPS | Volunteers were allowed to go home 24 hours post-LPS. | No safety concerns and dropouts indicated | USA |
| [2] Characterization of inflammation and immune  cell modulation induced by low-dose LPS  administration to healthy volunteers  [unique] | Showed the feasibility of low dose endotoxin administration and ex vivo endotoxin challenges in clinical studies | 24  LPS received by 6 volunteers each within 3 cohorts.  LPS = 18 | Healthy Caucasian male volunteers (non-smoking) | 18 – 28  Mean 23 | 0.5, 1, 2 ng/kg  U.S. Reference Escherichia Coli  CC-RE-Lot 3  O113:H, 10: K negative  NIH  Bethesda, Maryland | Electrocardiogram (ECG)  Blood pressure  Body temperature | IL-6, IL-8, IL-1β, and TNF-α | None | Increased heart rate and body temperature (dose-dependent).  Monocyte count decreased (dose-dependent).  Blood platelet count decreased  Increased neutrophil and leukocyte counts  Decreased lymphocyte count, erythrocyte count, and eosinophil count, basophil count and hemoglobin and hematocrit  Activated partial thromboplastin time (APTT) decreased  C-reactive protein increased (dose-dependent)  Increased IL-6, IL-8, and TNF-α (dose-dependent)  Levels of IL-1β increased (dose-dependent) | Blood samples were drawn up to 72 hours after LPS administration in vivo | The most commonly observed adverse events were feeling cold and headache.  No safety concerns and dropouts indicated | The Netherlands |
| [3] Amplified gut feelings under inflammation and depressed mood: A randomized fMRI trial on interoceptive pain in healthy volunteers  [unique] | Examined the effects of visceral pain stimuli in healthy individuals. | 39  LPS (n = 39)  Male (n = 22)  Female (n = 17) | Healthy volunteers | 20 – 39  29.5 | 0.4 ng/kg  Escherichia coli  serotype O113:H10  Lot H0K354  United States Pharmacopeia  Rockville, Maryland | Not mentioned | TNF-α and IL-6 | The pain levels of volunteers were assessed. | Increased cytokine levels  Increased sickness symptoms  Heightened the perception of visceral pain  Increased brain activity | Collection of blood sample was done 6 hours post-injection | 45 volunteers successfully finished all screening sessions despite a turnout of 63 healthy volunteers. However, 6 participants were disqualified.  No safety concerns and dropouts indicated (among the final 39 volunteers) | Germany |
| [4] First-in-Human Studies of MW01-6-189WH, a Brain-Penetrant, Antineuroinflammatory Small-Molecule Drug Candidate: Phase 1 Safety, Tolerability, Pharmacokinetic, and Pharmacodynamic Studies in Healthy Adult Volunteers  [unique] | Evaluated whether MW189, when administered to volunteers exposed to LPS can lead to alterations in plasma cytokine levels. | 18  LPS (n = 16) | Healthy male volunteers | 18 - 40  Mean 29 | 2 ng/kg  Escherichia coli LPS | ECG, and body temperature were measured. | TNF-α, IL-6, IL-8, IL-10, IL-1ra and CCL2 | The subjects reported symptoms following dosing. | Flu-like symptoms were exhibited  Cytokine levels increased | Up to 24 hours | No safety concerns and dropouts indicated. | USA |
| [5] B-cell dynamics during experimental endotoxemia in humans  [unique] | Studied the kinetics of the B-cell compartment, including regulatory B cells (Breg) in a human endotoxemia model. | 20  LPS (n = 20) | Healthy Caucasian males | 18.8 – 34.9  Mean 26.85 | 0.8 ng/kg  Eschericha coli LPS  LOT HOK354  U. S. Pharmacopeial Convention Inc.  Rockville, Maryland | Heart and respiratory rates, pulse oximetry using Kernmed Oled from Ettlingen, Germany, and blood pressure using Dinamap Compact T from Critikon, Norderstedt, Germany. | B-cell activating factor (BAFF), TNF-α and IL-10 | None | Increased heart rate  Increased respiratory rate  Increased body temperature  Decreased systolic pressure  Decreased lymphocyte counts  Decreased T-cell counts  Reduction in absolute CD19^+^ B cell counts  Memory B cells were moderately diminished  Breg experienced a decrease  BAFF increased | Blood samples were drawn up to 72 hours post-LPS | A family-related acute gastroenteritis situation prevented one of the participants from finishing the LPS condition, extending beyond the 72-hour point.  No safety concerns indicated | Germany |
| [6] Platelet P2Y_12_ inhibitors reduce systemic inflammation and its prothrombotic effects in an experimental human model  [unique] | Studied the outcome of P2Y_12_ inhibition in a human endotoxaemia model | 30  LPS (n = 30) | Healthy male volunteers | Not mentioned | 2 ng/kg  Escherichia coli  National Institutes of Health  U. S. | Not mentioned | IL-6, IL-8, IL-10, and TNF-α | None mentioned | Flu-like symptoms were experienced by the volunteers  CCL2 levels increased  Growth colony stimulating factor increased  hsCRP increased  Cytokine levels increased (TNF-α, IL-6, and IL-8)  Monocytes temporarily accumulated  Peak absorbance value of fibrin clot increased  Lysis area elevated  Clot formation closely packed (fibrin clot density elevated and diameter of the fibrin fiber reduced)  D-dimer increased | (could not estimate) | No safety concerns and dropouts indicated. | UK |
| [7] Effect of the antihepcidin Spiegelmer lexaptepid on inflammation-induced decrease in serum iron in humans  [unique] | Assessed the efficacy of lexaptepid in inhibiting the decline in serum iron levels in a human endotoxaemia model  Top of Form  Bottom of Form | 24  LPS (n = 24) | Healthy male volunteers | Not mentioned | 2 ng/kg  Purified standard reference  Escherichia coli O:113  Clinical Center Reference Endotoxin  National Institutes of Health  Bethesda, MD | Not mentioned | TNF- α, IL-6, IL-10, and IL-1 receptor antagonist | None | Plasma hepcidin levels increased initially, eventuated by a subsequent decrease  Serum iron levels increased (possibly due to the administration of lexaptepid) | - | No safety concerns and dropouts indicated. | The Netherlands |
| [8] β2-glycoprotein I: A novel component of innate immunity  [unique] | Showed that LPS can be inactivated by β_2_GPI | 23  LPS (n = 23) | Healthy male volunteers | 23.9 ± 0.7 years  23.9 | 4 ng/kg  E coli LPS  lot G  US Pharmacopeia | Blood pressure, oral temperature, and heart rate were monitored | TNF- α, IL-6, and IL-8 | None | Flu-like symptoms were exhibited under the influence of LPS | Blood was drawn up till 24 hours post LPS | No safety concerns and dropouts indicated | The Netherlands |
| [9] A novel model of common Toll-like receptor  4- and injury-induced transcriptional themes  in human leukocytes  [unique] | Investigated similarities in transcription patterns and common functional modules affected by TLR4 activation through endotoxin exposure and non-infectious injury occurring at the onset in trauma patients. | 15  LPS (n = 15) | Healthy adult volunteers  Male (n = 9)  Female (n = 5) | 18 – 36  27 | 2 ng/kg  NIH  Clinical Center Reference Endotoxin  CC-RE-Lot2 | Not mentioned | Not mentioned | None | Gene expression was altered in response to the endotoxin challenge | The duration of the study covers a 24-hour period. | No safety concerns and dropouts indicated | USA |
| [10] A randomised trial on the effect of anti-platelet therapy on the systemic inflammatory response in human endotoxaemia  [unique] | Used the in vivo endotoxaemia model to study the effects of combined antiplatelet therapies on systemic inflammation | 40  Ticagrelor and ASA (n = 10)  Clopidogrel and ASA (n = 10)  Placebo and ASA (n = 10)  Placebo only (n = 10)  LPS (n = 40) | Healthy male volunteers | 18 – 35  26.5 | 1 ng/kg + 1 ng/kg/hr for 3 hours  Purified lipopolysaccharide  U.S. Standard Reference Endotoxin  Escherichia Coli O:113  Pharmaceutical Development Section  National Institutes of Health  Bethesda, MD | Heart rate, temperature, and blood pressure were monitored | TNF-α, IL-6, IL-8, IL-10 and IL-1RA | None | Increased plasma concentration of all measured cytokines as well as MCP-1, MIP-1α and MIP-1β  Decreased mean arterial blood pressure (MABP) and increased both heart rate and body temperature.  Elevated levels of plasma adenosine  (LPS was administered on day 7^th^ day of the trial. Subject were already on treatment; these results may not be due to LPS only) | LPS was administered on the 7^th^ day. | No safety concerns and dropouts indicated  It was indicated that within the clopidogrel-ASA and ticagrelor-ASA groups, one participant reported experiencing a hematoma and the other experienced mild shortness of breath, while still maintaining the ability to tolerate exercise. | The Netherlands |
| [11] A randomized double-blind, placebo-controlled clinical phase IIa trial on safety, immunomodulatory effects and pharmacokinetics of EA-230 during experimental human endotoxaemia  [unique] | Evaluated the safety of EA-230 with the aid of the human endotoxaemia model | 36  Placebo (n = 12)  15 mg/kg/h EA-230 (n = 8)  45 mg/kg/h EA-230 (n = 8)  90 mg/kg/h EA-230 (n = 8)  LPS (n = 36) | Healthy adult males | Placebo (23 ± 3)  15 mg/kg/h EA-230 (22 ± 1)  45 mg/kg/h EA-230 (22 ± 3)  90 mg/kg/h EA-230 (22 ± 2)  EA-230? Reported in the article as β‐hCG (Beta-human chorionic gonadotropin) ‐ derived immunomodulatory tetrapeptide  22 | 2 ng/kg  Purified LPS  U.S. Standard Reference Endotoxin  Escherichia coli O:113  Pharmaceutical Development  National Institutes of Health  Bethesda, MD | Heart rate (3-lead ECG), blood pressure (Philips MP50 patient monitor), and temperature (infrared tympanic thermometer (First‐Temp Genius 2; Covidien, Dublin, Ireland)) | IL-6, IL‐10 and TNF‐α, IL‐8, (MCP)‐1, IL‐1 (RA), (MIP)‐1α and MIP‐1β | LPS induced sickness symptoms were scored according to their severity were considered. | Endotoxaemia resulted in flu-like symptoms.  Increased body temperature.  Mean arterial blood pressure decreased.  Increased heart rate.  EA-230 attenuated sickness behaviors induced by LPS in vivo. | Physiological monitoring was done till 8 hours post LPS.  Volunteers came back for follow-ups till the 14^th^ day after drug administration. | No safety concerns and dropouts indicated | The Netherlands |
| [12] Adenosine infusion attenuates soluble RAGE in endotoxin-induced inflammation in human volunteers  [unique] | Investigated the anti-inflammatory effects of adenosine in the human endotoxaemia model. | 16  LPS (n = 16) | Healthy male volunteers | 27.1 ± 7  27.1 | 2 ng/kg  National Reference Endotoxin  E.Coli Lot.G 1 and Lot G2B274  U. S. Pharmacopeial Convention  Rockville, MD | Body temperature, heart rate, and blood pressure | IL-6, IL-10, and TNF- α | All volunteers reported discomfort and fatigue | Flu-like symptoms were exhibited  Heart rate increased  Body temperature increased  MAP increased  Level of cytokines increased  Resistin was elevated | Samples were collected for 5 hours after the administration of LPS.  NO was examined for 5 hours after the administration of LPS.  Urine samples were taken 11–13 hours after the administration of LPS. | No safety concerns and dropouts indicated | Sweden |
| [13] Altered temporal variance and functional connectivity of BOLD signal is associated with state anxiety during acute systemic inflammation  [unique] | Investigated changes in the anxiety and mood among volunteers administered with LPS and the neural processes involved in changes in state anxiety. | 43  LPS (n = 20) | Male volunteers (healthy) | 18 – 45  31.5 | 0.4 ng/kg  Reference Standard Endotoxin  lot G3E069  U. S Pharmacopeia  Rockville, MD | Heart rate, body temperature, and blood pressure | IL-6, and TNF-α | Researchers assessed mood and anxiety symptoms  “The State Trait Anxiety Inventory (STAI)” was used for the assessment of anxiety  “The Multidimensional Mood Questionnaire (MDBF)” was used for the assessment of mood | IL-6 and TNF- α increased  The LPS group reported higher anxiety scores.  The positive-mood scores in the LPS group decreased | The assessment of the mood and anxiety symptoms lasted for a period of 6 hours post-LPS | Some participants were excluded from certain measurements. The reason for exclusion was not indicated as that caused by LPS injection.  No safety concerns | Germany |
| [14] An fMRI study of cytokine-induced depressed mood and social pain: The role of sex differences  [unique] | Investigated the effects of cytokine activation on neural systems and depressed mood in the human endotoxaemia model | 39  LPS (n = 20)  3 volunteers received LPS but were not scanned (no participation in the neuroimaging session) | Healthy volunteers | 18 – 36  27 | 0.8 ng/kg  E. coli group O:113  National Institutes of Health  Clinical Center reference endotoxin | Temperature, pulse rate, and blood pressure | IL-6 | Assessment of depressed mood and physical symptoms by participants | IL-6 increased  Cortisol levels increased  Physical symptoms self-reportedly increased  Pulse and temperature increased  Depressed mood self-reportedly increased | The scanning session was completed approximately 2 hours after the injection of LPS. | No safety concerns and dropouts indicated | USA |
| [15] Anterior insula morphology and vulnerability to psychopathology-related  symptoms in response to acute inflammation  [unique; but described elsewhere]  [unique] | Investigated the link between brain morphological measurement and the subjective reactions to stimulated inflammatory responses. | 52  LPS (n = 28) | Healthy volunteers | 18 – 50  (28.6 ± 7.1)  28.6 | 0.6 ng/kg  E. coli  Lot G3E0609  U.S Pharmacopeia  Rockville, MD | None | TNF-α and IL-6, and IL-8 | The Sickness Questionnaire (SicknessQ) scores was used to assess subjective symptoms  The State Trait Anxiety Inventory (STAI-S)” was used for the assessment of state anxiety | State anxiety and sickness symptom increased  IL-6 and TNF-α increased | Blood samples were drawn up to 5 hours post-LPS | Cytokine and self-reported questionnaire data were missing for 3 individuals. 31 participants were assigned to the LPS group, however, only data from 28 of them were used  No safety concerns and dropouts indicated | Sweden |
| [16] Antibiotic-induced gut microbiota disruption during human endotoxemia: a randomised controlled study  [unique] | Used the human endotoxaemia model to investigate whether alterations in the gut microbiota influence inflammation | 16  LPS (n = 16) | Healthy young males (Caucasian) | 18 – 25  21.5 | 2 ng/kg  E. coli O113  Reference Endotoxin  CC-RE lot 3  National Institutes of Health  Bethesda, Maryland | Temperature, heart rate, and blood pressure | TNF-α, IL-1β, IL-6, IL-8, IL-10, and IL-12p70 | None | LPS injection resulted in flu-like symptoms (headache, fever, chills and nausea)  Cytokine levels increased  Transient increase in heart rate, blood pressure, and temperature  The number of neutrophils elevated  Blood leucocyte counts increased  Plasma concentrations of Myeloperoxidase (MPO) increased | Blood samples were collected up to 8 hours post-LPS | No safety concerns and dropouts indicated | The Netherlands |
| [17] APOε4 is associated with enhanced in vivo innate immune responses in human subjects  [unique] | Investigated the relationship between *APOε4* genotype and inflammatory responses in the human endotoxaemia model | 35  LPS (n = 35)  Male (n = 22)  Female (n = 13)  European American (n = 14)  Asian (n = 9)  African American (n = 8)  Hispanic (n = 4)  APOε3/APOε3 genotype (n = 18)  APOε3/APOε4 genotype (n = 7) | Healthy volunteers | 18 - 40  24.5 ± 1.0  24.5 | 2 ng/kg  LPS (CC-RE, lot 2) | Temperature | TNF-α and IL-6 | None | Patients with APOε3/APOε4 genotype experienced an increase in body temperature as compared to APOε3/APOε3 patients  Patients with APOε3/APOε4 genotype experienced elevated TNF- α levels as compared to APOε3/APOε3 patients  Increased temperature and TNF-α levels seen in patients with ε4^+^ | Blood samples were collected up to 24 hours post-LPS | No safety concerns and dropouts indicated | USA |
| [18] Alterations in functional connectivity of resting state networks during experimental endotoxemia – An exploratory study in healthy men  [unique] | Investigated the potential impact of LPS on the functional connectivity between different brain regions | 45  LPS (n = 20) | Healthy male volunteers | 18 – 45  26.2 ± 0.5  26.2 | 0.4 ng/kg  Reference Standard Endotoxin  lot G3E069  United States Pharmacopeia  Rockville, MD | Temperature, heart rate, and blood pressure | TNF-α and IL-6 | Self-evaluated alertness and positive mood | Leukocyte counts increased  TNF-α increased  IL-6 increased  Cortisol increased  Body temperature increased  Self-evaluated alertness and positive mood decreased | Blood samples were drawn up to 6 hours post-LPS | No safety concerns and dropouts indicated | Germany |
| [19] Blunting the response to endotoxin in healthy subjects: effects of various doses of intravenous fish oil  [unique] | Investigated the impact of multiple doses of intravenously administered fish oil (FO) in a human endotoxemia model | 23  LPS (n = 23) | Healthy male volunteers | 18 – 35  26.5 | 2 ng/kg  E. Coli  USP  Rockville, MD  batch no. 2 | Heart rate  Respiratory rate  Rectal temperature (measured with Hellige Servomed)  Non-invasive arterial blood pressure (monitored using Critikon Dinamap)  Cardiac output (assessed with thoracic bioimpedance using NCCOM3 cardiodynamic monitor by BoMed, Irvine, California)  Oxygen saturation (measured with Ohmeda Biox 3740 Pulse oximeter) | TNF-α | Headache, nausea, vomiting, and muscle pain | Heart rate increased  Cardiac output increased  Body temperature increased  Sickness symptoms were exhibited  TNF-α increased  Norepinephrine increased  Epinephrine increased  Adrenocorticotropic hormone (ACTH) levels increased | Blood sample was drawn over an 8-hour period after the LPS challenge | No safety concerns and dropouts indicated | Switzerland |
| [20] Safety, tolerability and pharmacokinetics/ pharmacodynamics of the adrenomedullin antibody adrecizumab in a first-in-human study and during experimental human endotoxaemia in healthy subjects  [unique] | Adrecizumab’s safety and other characteristics were studied for the first time in man and second in a human endotoxaemia model | 48  Endotoxemia (n = 24) | Healthy male volunteers | 18 – 35  26.5 | 1 ng/kg, then 1 ng/kg/h for 3 hours  Escherichia coli  type O113 LPS  List Biological Laboratories Inc.  Campbell, CA | Heart rate, blood pressure and peripheral oxygen saturation were monitored using a Philips MP50 patient monitor (Philips, located at Eindhoven, the Netherlands)  Temperature was measured using the FirstTemp Genius 2 device from Sherwood Medical in St Louis, Missouri, USA, and the 12-lead ECG was recorded using the Philips PageWriter Trim II, from Philips in Amsterdam, the Netherlands | TNF-α, interleukin-6 (IL-6), interleukin-8 (IL-8), interleukin-10 (IL-10), granulocyte-colony stimulating factor, interferon gamma-induced protein 10 and monocyte chemoattractant protein 1 | Symptom scores were assessed | Body temperature increased  Heart rate increased  Mean arterial pressure decreased  Symptom scores increased  Cytokine levels increased  Chemokines increased | They were released 8 hours post-LPS | A severe adverse event was documented (participant was diagnosed with type 1 diabetes mellitus). Not LPS related and possibly had it developing before the study.  No safety concerns and dropouts indicated | The Netherlands |
| [21] C1-esterase inhibitor attenuates the inflammatory response during human endotoxemia  [unique] | Investigated the impact of C1-esterase inhibitor on immune response in an experimental human endotoxemia model | 20  LPS (n = 20) | Healthy male volunteers | 22.1 ± 3.1  22.1 | 2 ng/kg  U.S. Standard Reference Endotoxin  Escherichia coli O:113  Pharmaceutical Development Section  National Institutes of Health  Bethesda, MD | Heart rate measured with a five-lead ECG  Blood pressure  Body temperature (measured with FirstTemp Genius; Tyco Healthcare, located at Hampshire, United Kingdom) | TNF-α, IL-10, monocyte chemotactic protein-1 (MCP-1), IL-1β, and IL-1RA | Symptom scores were assessed | Sickness symptom exhibited (headache)  Sickness symptom scores increased  Blood pressure decreased  Heart frequency increased  Body temperature increased  Cytokine levels increased  Indicators of endothelial activation increased | Data was collected up to 24 hours post-LPS | No safety concerns and dropouts indicated | The Netherlands |
| [22] Challenging the challenge: A randomized controlled trial evaluating the inflammatory response and pain perception of healthy volunteers after single-dose LPS administration, as a potential model for inflammatory pain in early-phase drug development  [unique] | Examined how healthy volunteers perceive pain following the administration of LPS | 24  LPS (n = 24) | Healthy male volunteers | 19 – 52  35.5 | 1 ng/kg (n = 12) and 2 ng/kg (n = 12)  E. Coli-purified LPS  GMP-grade  Lot#94332B4  List Biological Laboratories Inc.  CA | Temperature | IL-6, TNF-α, IL-8, IL-1ra, IL-10, IL-1b | Pain levels were assessed | Cytokine levels increased  C-reactive protein concentrations increased  Cortisol levels increased | Took 7-21 days between test occasions. | Two volunteers were replaced (not reported as a result of the administration of LPS)  No safety concerns indicated | The Netherlands |
| [23] Circulating adenosine increases during human experimental endotoxemia but blockade of its receptor does not influence the immune response and subsequent organ injury  [unique] | Investigated the effect of the administration of LPS on circulating adenosine concentration. | 43  LPS (n = 30) | Healthy male volunteers | 21 – 25  23 | 2 ng/kg  U.S. Reference E. coli endotoxin  Escheria coli O:113  Clinical Center Reference Endotoxin  National Institutes of Health  Bethesda, MD | Blood pressure, body temperature (measured with an infrared tympanic thermometer from Sherwood Medical, located at ‘s-Hertogenbosch, The Netherlands), and heart rate (ECG) | TNF-α, IL-6, IL1RA, and IL-10 | Symptom scores were taken | Sickness symptoms were observed  Body temperature increased  White blood cell counts decreased initially, then increased later  Blood pressure decreased  Heart rate increased  Elevated forearm blood flow  Cytokine levels increased | Physiological measurement done up to 8 hours post-LPS | No safety concerns and dropouts indicated | The Netherlands |
| [24] Colistin Reduces LPS-Triggered Inflammation in a Human Sepsis Model In Vivo: A Randomized Controlled Trial  [unique] | Investigated the effect of colistin on inflammatory response in humans | 15  LPS (n = 15) | Healthy male volunteers | 19 – 40  29.5 | 2 ng/kg  E. coli 0113  Reference Endotoxin  CC-RE Lot 3  National Institutes of Health  Bethesda, MD | Noninvasive arterial blood pressure, sublingual body temperature, and heart rate | IL-6, IL-8, IL-1β, TNF-α | Self-reported symptoms (joint pain, headache, and shivering) | Flu-like symptoms were experienced  Colistin affected inflammatory response due to LPS; reduced cytokine response | Up to 24 hours | After the administration of lipopolysaccharide (LPS), two volunteers who had received a prior saline placebo experienced a temporary absence of heartbeat (asystole), accompanied by a temporary loss of consciousness that subsequently resolved.  One volunteer did not complete all the study periods (not as a result of LPS) | Austria |
| [25] Common studied polymorphisms do not affect plasma cytokine levels upon endotoxin exposure in humans  [unique] | Investigated the relationship between SNPs and LPS-induced cytokine response in a human endotoxaemia model | 200  LPS (n = 200) | Healthy male volunteers (Caucasian) | 25 ± 4  25 | 0.1 ng/kg  E. coli endotoxin  lot G2B274  United States Pharmacopeial Convention  Rockville, Maryland | Not mentioned | IL-6, IL-10, IL-18, and TNF-α | None | TNF-α and IL-6 levels increased | Blood samples were drawn up to 3 hours | No safety concerns and dropouts indicated | Denmark |
| [26] Continuous Administration of Enteral Lipid- and Protein-Rich Nutrition Limits Inflammation in a Human Endotoxemia Model  [unique] | Investigated the potential immunomodulatory effects of a specially formulated enteral nutrition in a human endotoxaemia model | 18  12 from this main study, 6 from another  LPS (n = 12) | Healthy male volunteers | - | 2 ng/kg  U.S. reference endotoxin  Escherichia coli O:113  Clinical Center Reference Endotoxin  Lot EC-5  National Institute of Health  Bethesda, MD | Heart rate (3-lead electrocardiogram)  Body temperature recorded with an infrared tympanic thermometer (from FirstTemp Genius; Sherwood Medical, located at Crawley/ Sussex, UK)) | TNF-α, IL-1RAs, IL- 6, and IL-10 | Symptom scores were taken | Mean arterial blood pressure decreased  Heart rate increased  Body temperature increased  White blood cell counts increased  Symptom scores increased | Blood collected from volunteers up to 24 hours | No safety concerns and dropouts indicated | The Netherlands |
| [27] CytoSorb hemoperfusion markedly attenuates circulating cytokine concentrations during systemic inflammation in humans in vivo  [unique] | Studied the influence of CytoSorb hemoperfusion on cytokine levels in a repeated in vivo model of LPS injection | 24  LPS (n = 24) | Healthy male volunteers | 18 – 35  26.5 | 1 ng/kg + 0.5 ng/kg/hr  E. coli type O:113  lot no. 94332B1  List Biological Laboratories  Campbell, U.S.A | Intra-arterial blood pressure and heart rate (monitored with a 4-lead ECG; M50 Monitor, from Philips, located at Eindhoven, the Netherlands)  Radial artery pressure transducer (from Edwards Lifesciences, located at Irvine, California, USA)  Body temperature was monitored with a tympanic thermometer (from FirstTemp Genius 2, Covidien, located at Dublin, Ireland) | TNF, IL-6, monocyte chemoattractant protein (MCP)-1, IL-8, IL-10, granulocyte colony stimulating factor (G-CSF), macrophage inflammatory protein (MIP)-1α, and interferon-γ-induced protein (IP)-10 | Symptom scores were taken | Cytokine levels increased  Blood pressure decreased  Heart rate increased  Body temperature increased  Symptom scores increased | Blood samples collected up to 8 hours post-LPS | No safety concerns and dropouts indicated | The Netherlands |
| [28] Defibrotide enhances fibrinolysis in human endotoxemia – a  randomized, double blind,  crossover trial in healthy volunteers  [unique] | To investigate the effects of Defibrotide on inflammatory response in a human endotoxaemia model | 20  LPS (n =16) | Healthy volunteers  Males (n = 18)  Females (n = 2) | More than 18 years (volunteers ≥ 18)  - | 2 ng/kg | Vital signs were taken as reported in the article. None of them were mentioned | TNF-α, and IL-6 | Volunteers reported symptoms | Thrombin generation increased in vivo  Blood clotting time was reduced, as confirmed by thromboelastometry  Fibrinolytic system was triggered  Leukocyte counts increased  Flu-like symptoms were experienced by the volunteers | Blood samples were collected up to 24 hours post-LPS | No safety concerns and dropouts indicated in the LPS group | Austria |
| [29] Development of endotoxin tolerance does not influence the response to a challenge with the mucosal live-attenuated influenza vaccine in  humans in vivo  [unique] | Investigated the impact of an immune response triggered by bacterial infection and the development of tolerance to endotoxin on the reaction to a further infection with influenza | 30  LPS (n = 15) | Healthy male volunteers | 18 – 35  26.5 | 2 ng/kg  US Standard Reference Endotoxin  Escherichia coli O:113  Pharmaceutical  Development Section  National Institutes of Health  Bethesda, Maryland | Heart rate was measured with a 3-lead ECG  Intra-arterial blood pressure (measured with a Philips MP50 patient monitor from Philips, located at Eindhoven, The Netherlands)  An infrared tympanic thermometer was used to measure temperature (FirstTemp Genius 2 instrument, Sherwood Medical, located at Crawley/Sussex, United Kingdom) | TNF-α, IL-6, IL-10, IL-8, G-CSF, IFN-γ, IL-1β, IL-13, IP-10, IFN- α, IFN-β | Symptom scores were assessed | TNF-α, IL-6, and IL-10 levels increased  Decreased blood pressure  Increased heart rate  Increased body temperature  Elevated count of neutrophils  Reduced monocyte count  Reduced number of lymphocytes | Blood samples were drawn up to 8 hors post-LPS | No safety concerns  2 volunteers were excluded (not as a result of LPS) | The Netherlands |
| [30] Differential dose-dependent effects of prednisolone on shedding of endothelial adhesion molecules during human endotoxemia  [unique] | Studied how the levels of soluble adhesion molecules are influenced when healthy volunteers are exposed to increasing doses of prednisolone in a human endotoxaemia model | 32  LPS (n = 32) | Healthy male volunteers | 23.9 ± 0.7  23.9 | 4 ng/kg  Escherichia coli  lot G  U.S. Pharmacopeia  Rockville, MD | None | None | None | sE-selectin levels increased  Soluble intercellular adhesion molecule-1 (sICAM-1) levels increased  soluble vascular cell adhesion molecule-1 (sVCAM-1) levels increased | Blood samples were collected up to 24 hours post-LPS | No safety concerns and dropouts indicated | The Netherlands |
| [31] Dipyridamole augments the antiinflammatory response during human endotoxemia  [unique] | Investigated the effects of dipyridamole in a human endotoxaemia model | 20  LPS (n = 20) | Healthy male volunteers | Placebo (21.4 ± 1.8)  Dipyridamole (22 ± 2.6)  21.7 | 2 ng/kg  Escherichia coli O:113  Clinical Center Reference Endotoxin  National Institutes of Health  Bethesda, MD | Blood pressure, body temperature, and heart rate | TNF-α, IL-6, IL-1RA, and IL-10 | None | Decreased white blood cell count initially, then increased later  Increased circulating monocytes  Body temperature increased  Flu-like symptoms experienced  Blood pressure decreased  Heart rate increased  Fore-arm blood flow increased  ICAM levels increased  VCAM levels increased  Total antioxidant capacity elevated | Up to 24 hours | No safety concerns and dropouts indicated | The Netherlands |
| [32] Dobutamine does not influence inflammatory pathways during human endotoxemia  [unique] | Investigated the effects of dobutamine in a human endotoxaemia model | 16  LPS (n = 16) | Healthy male volunteers | 25 ± 2  25 | 4 ng/kg  Escherichia coli  Lot G  UPS  Rockville, MD | Body temperature  Heart rate and blood pressure (Dinamap) | IL12p70, TNF- α, IL-6, IL-8, IL- 10, and IL-1β | Symptom scores were recorded | Body temperature increased  Flu-like symptoms were experienced  TNF-α, IL-6, and IL-8 levels increased  The levels of soluble tissue factor in the plasma increased  Levels of prothrombin fragment F1+2 elevated  The levels of thrombin-antithrombin complexes increased  The levels of soluble E selectin in the plasma increased  Plasma levels of secretory phospholipase A_2_ (PLA_2_) increased  The number of neutrophils decreased momentarily, and increased later | Blood samples were drawn up to 22 hours post-LPS | No safety concerns and dropouts indicated | The Netherlands |
| [33] Duffy antigen modifies the chemokine response in human endotoxemia  [unique] | Investigated how Duffy antigen impacts inflammation in a human endotoxaemia model | 32  LPS (n = 32) | Healthy male volunteers | 23 – 31  22 | 2 ng/kg  National Reference Endotoxin  Escherichia coli  CC-RE-Lot 2  National Institutes of Health  Bethesda, Maryland | Oxygen saturation, heart rate, ECG, and blood pressure | TNF, IL-6, IL-10, (MCP)-1, growth-related oncogene (GRO)-α, G-CSF, MIP-1β | None | Cytokines (IL-6 and IL-10) levels increased  Whole blood GRO- α increased  MCP-1 increased  Interleukin-8 mRNA increased  Neutrophilia  Monocytopenia  Lymphopenia | Blood samples were drawn up to 24 hours post-LPS | No safety concerns and dropouts indicated | Austria |
| [34] E. coli Endotoxin Modulates the Expression of Sirtuin Proteins in PBMC in Humans  [unique] | Investigated the expression profiles of Sirtuins in peripheral blood mononuclear cells (PBMC) in a human endotoxaemia model | 20  LPS (n = 20) | Healthy male volunteers | 19 – 40  29.5 | 2 ng/kg  E. coli  US Standard Reference Endotoxin  NIH-CC  Bethesda, Maryland | Body temperature | None | None | Flu-like symptoms were experienced  Body temperature increased  Leukocyte cell counts elevated  The mRNA expressions of SIRT1 and SIRT3 in human peripheral blood mononuclear cells (PBMC) were decreased | Blood samples were drawn up to 24 hours post-LPS | No safety concerns and dropouts indicated | Austria |
| [35] Effect of Hypoglycemia on Inflammatory Responses and the Response to Low-Dose Endotoxemia in Humans  [unique] | Investigated the impacts of Hypoglycemia on inflammatory responses, and in a human endotoxaemia model | 24  LPS (n = 24) | Healthy volunteers | 19 – 26  22.5 | 0.3 ng/kg  Escherichia coli O:113  Clinical Centre Reference Endotoxin  National Institutes of Health  Bethesda, Maryland | Blood pressure | None | None | White blood cells count increased  Lymphocyte counts reduced  Monocyte count decreased momentarily, then it was restored afterwards  Heightened the release of cortisol  Reduced the expression of CX_3_CR1  CX_3_C chemokine ligand 1 concentrations elevated  Elevated the expression of CD11b | Blood samples were drawn up to 6 hours post-LPS | No safety concerns and dropouts indicated | UK |
| [36] Effect of systemic high dose vitamin C therapy on forearm blood flow reactivity during endotoxemia in healthy human subjects  [unique] | Investigated the effect of the administration of high systemic doses of vitamin C on forearm blood circulation reactivity in a human endotoxaemia model | 36  LPS (n = 36) | Healthy male volunteers | 27 ± 6  27 | 20 IU/kg  National Reference Endotoxin  National Institutes of Health (NIH) | Blood pressure  Pulse rate  ECG  Tympanic temperature | TNF-α | None | Flu-like symptoms were experienced  Body temperature increased  TNF-α increased  Decreased levels of vitamin C in the plasma  Reduced the acetylcholine (ACh)-dependent augmentation in forearm blood flow (FBF) | (could not estimate) | No safety concerns and dropouts indicated | Austria |
| [37] Effect of Vasopressors on the Macro- and Microcirculation during Systemic Inflammation in Humans in Vivo  [unique] | Assessed the impact of three vasopressor agents on two concepts, microcirculation and systemic blood flow dynamics, in a human endotoxaemia model | 40  LPS (n = 40) | Healthy male volunteers | 18 – 35  26.5 | 2 ng/kg  E. coli LPS | Blood pressure parameters  Heart rate  Cardiac output (CO) | None | None | Flu-like symptoms were experienced  Blood pressure parameters decreased  Heart rate increased  Cardiac output increased  Systemic vascular resistance (SVR) decreased  Microvascular density and blood flow reduced | (could not estimate) | No safety concerns and dropouts indicated | The Netherlands |
| [38] Effects of acute systemic inflammation on  the interplay between sad mood and affective cognition  [unique] | Studied how acute inflammation and a sad mood interact and influence cognitive function; mood-related, in a human endotoxaemia model | 15  LPS (n = 15) | Healthy male volunteers | 18 – 40  29 | 0.8 ng/kg  Reference standard endotoxin  Escherichia coli  serotype O113:H10:K- negative  lot H0K354  United States Pharmacopeia  Rockville, MD, USA | Blood pressure (measured with a Dinamap Compact T device, from Critikon, Located at Norderstedt, Germany)  Body temperature (measured using an intraaurical thermometer)  Heart rate (utilized pulse oximetry; with a Kernmed Oled device, from Ettlingen, Germany) | IL-6 and TNF-α | Assessed changes in symptoms and mood reported by volunteers | White blood cell counts increased  TNF-α levels increased  IL-6 levels increased  Body temperature increased  Dysthymia, as reported by the volunteers, increased  Euthymia, as reported by the volunteers, decreased  Symptom scores increased | Blood was drawn up to 6 hours post-LPS | No safety concerns and dropouts indicated | Germany |
| [39] Effects of fish oil on the neuro-endocrine responses to an endotoxin challenge in healthy volunteers  [unique] | Investigated the impact of orally administered fish oil supplements in a human endotoxaemia model | 15  (16 were recruited)  LPS (n = 15) | Healthy male volunteers | 26 ± 3.1  26 | 2 ng/kg  Escherichia coli  USP  Rockville,MD | Rectal temperature (measured with Hellige, Servomed)  Heart rate  Cardiac output was measured (used NCCOM3 cardiodynamic monitor from BoMed, located at Irvine, California) through a bioimpedance process  Arterial blood pressure (non-invasively measured)  Respiratory rate  Oxygen saturation (measured with a pulse oximeter device; Ohmeda, Biox 3740) | TNF-α and IL-6 | Volunteers reported symptoms (muscle ache, nausea, and headache) | Flu-like symptoms were experienced  Heart rate increased  Cardiac output increased  Body temperature increased  TNF-α and IL-6 levels increased  ACTH increased  Norepinephrine increased  Epinephrine increased  CRP increased  (Energy expenditure) EE increased  Glucose in plasma reduced initially, and increased afterwards  Reduced net glucose oxidation  Elevated net fat oxidation  Elevated free fatty acids | - | The research protocol was reportedly stopped by a volunteer  Two volunteers vomited  No safety concerns | Switzerland |
| [40] Effects of low dose endotoxemia on endothelial progenitor cells in humans  [unique] | Investigated the effects of LPS infusion on EPC levels | 36  LPS (n = 32) | Healthy male volunteers | IQR (22 -29)  Median age (26)  25.5 | 2 ng/kg  National Reference Endotoxin  Escherichia coli  CC-RE-Lot 2  National Institutes of Health  Bethesda | Heart rate, ECG, blood pressure and oxygen saturation were measured with a Care View System, from Hewlett Packard located at Böblingen, in Germany | TNF-α | None | Flu-like symptoms were experienced  Lymphocyte counts decreased  Circulating endothelial progenitor cells (EPCs) decreased  Colony-Forming Units CFUs decreased  Vascular endothelial growth factor (VEGF) concentrations increased  Granulocyte colony-stimulating factor (G-CSF) increased  TNF-α increased | Up to 24 hours | No safety concerns and dropouts indicated | Austria |
| [41] Emotional expressions of the sick face  [unique] | Investigated the changes in facial expression of emotion in a human endotoxaemia model | 22 | Healthy volunteers | 18 – 50  34 | 2 ng/kg  Escherichia coli endotoxin  Lot HOK354  CAT number 1235503  United States Pharmacopeia  Rockville, Maryland | Tympanic temperature | TNF-α, IL-6, and IL-8 | Assessed sickness ratings | Changed facial expression of emotion | (could not estimate) | No safety concerns  4 dropouts indicated (not due to LPS) | Sweden |
| [42] Endotoxemia causes central downregulation of sympathetic vasomotor tone in healthy humans  [unique] | Studied how LPS affects the activity of sympathetic nerves involved in blood vessel constriction and the general control of blood pressure | 18  LPS (n = 11) | Healthy male volunteers | LPS (28.7 ± 6.6)  28.7 | 4 ng/kg  Escherichia coli O:113  U.S. Standard Reference Endotoxin  Lot G-1  Food and Drug Administration  Bethesda, Maryland | Oscillometric Blood pressure (measured with a Welch Allyn Tycos device)  Heart rate  Heart rate variability  Muscle sympathetic nerve activity (MSNA) | TNF-α and IL-6 | None | Flu-like symptoms were experienced  Body temperature increased  Cytokine levels increased  Epinephrine levels increased  Norepinephrine levels increased | Blood was drawn up to 3 hours post-LPS | No safety concerns  Some data from volunteers were excluded (not reported as due to LPS). Out of the 11 volunteers in the LPS group, 7 had their recordings considered for analysis. | Germany |
| [43] Erythropoietin augments the cytokine response to acute endotoxin-induced inflammation in humans  [unique] | Investigated the anti-inflammatory outcomes of erythropoietin (EPO) in a human endotoxaemia model | 26  LPS (n = 16) | Healthy male volunteers | LPS (26 ± 2)  26 | 0.1 ng/kg  E. coli endotoxin  batch G2 B274  U. S. Pharmacopeial Convention, Inc.  Rockville, Maryland | Blood pressure (non-invasive)  Temperature  Heart rate | TNF-α, IL-6 and IL-10 | None | Heart rate reduced (among the groups)  Temperature increased  Cytokine levels increased  Lymphocyte counts reduced  Neutrophil counts elevated | Blood samples were collected up to 3 hours post-LPS | No safety concerns and dropouts indicated | Denmark |
| [44] Experimental endotoxemia induces adipose inflammation and insulin resistance in humans  [unique] | Investigated the effects of LPS administration into healthy human volunteers on the inflammation within adipose tissue and resistance of insulin | 20  LPS (n = 20) | Healthy volunteers | (27.3 ± 4.8)  18 – 40  27.3 | 3 ng/kg  U.S. standard reference endotoxin  lot no. CC-RE-LOT-1 2  Clinical Center, Pharmacy Department  NIH | Temperature  Heart rate | TNF-α, IL-6, MCP-1, and C-X-C motif ligand 10 (CXCL10) | None | Temperature increased  Heart rate increased  WBC counts increased  Cytokine levels increased  Resistin increased  Leptin increased  The ratio of Leptin-soluble leptin elevated  hsCRP increased  Cortisol increased  Free fatty acids increased  Growth hormone increased  mRNA concentrations of IL-6 elevated  mRNA concentrations of MCP-1 were triggered  mRNA concentrations of resistin (in the adipose tissue) increased  Certain Suppressor of cytokine signaling (SOCS) proteins were triggered | Up to 60 hours (endotoxaemia protocol) | No safety concerns and dropouts indicated | USA |
| [45] Factor V Leiden mutation enhances fibrin formation and dissolution in vivo in a human endotoxemia model  [unique] | Investigated Factor V Leiden’s processes in a human endotoxaemia model | 29  LPS (n = 29) | Healthy male volunteers | 20 – 69  44.5 | 2 ng/kg  National Reference Endotoxin,  E coli  The United States Pharmacopoial Convention Inc | None mentioned | None | None | Soluble fibrin elevated in factor V Leiden (FVL) volunteers  Peptidyl-prolyl cis-trans isomerase C (PPIC) increased  D-dimer antigen concentrations elevated  Fibrin degradation products (FDP) concentrations elevated in plasma | Blood samples were collected up to 24 hours | No safety concerns and dropouts indicated | Germany |
| [46] Frontline Science: Endotoxin-induced immunotolerance is associated with loss of monocyte metabolic plasticity and reduction of oxidative burst  Second study (n = 12)  LPS (n = 8) [unique] | Investigated the relationship between metabolic alterations and antimicrobial functions of monocytes in a human endotoxaemia model. | Second study (n = 12)   - LPS (n = 8) [unique] | Healthy male volunteers | 18 – 35  26.5 | 2 ng/kg  U.S. Standard Reference Endotoxin  E. coli O:113  Pharmaceutical Development Section  National Institutes of Health  Bethesda, Maryland | None mentioned | TNF-α, IL-6, IL-10, IL-1Ra, and IL-1β | None | Induced immunotolerance | Blood samples were drawn up to 7 days post-LPS | No safety concerns and dropouts indicated | The Netherlands |
| [47] Gamma-Irradiated Bacille Calmette-Guérin Vaccination Does Not Modulate the Innate Immune Response during Experimental Human Endotoxemia in Adult Males  [unique] | Investigated the BCG vaccine’s effect on innate immune system in a human endotoxemia model | 20  LPS (n = 20) | Healthy male volunteers | Placebo (n = 10)   - (19.0 – 24.3)   γ-irradiated BCG (n = 10)   - (19.8 – 22.0)   19.4 | 1 ng/kg  Escherichia coli O:113  Clinical Center Reference Endotoxin,  National Institutes of Health (NIH)  Bethesda, Maryland | Three-lead ECG measured heart rate  Blood pressure  Respiratory rate  Pulse oximetry device measured oxygen saturation  (Patient monitor from Philips – MP50 used for the measurements) | MCP-1, TNF-α, IFN-γ, IL-6, IL-8, IL-10, IL-1Ra, and IL-1β | Symptom scores were assessed | Heart rate increased  Flu-like symptoms experienced by volunteers  MAP decreased  Cytokine levels (MCP-1, TNF-α, IL-6, IL-8, IL-10 and IL-1RA) increased  White blood cells count temporary increased | Blood samples were collected up to 30 days post-experiment | No safety concerns and dropouts indicated | The Netherlands |
| [48] Hyperglycemia enhances coagulation and reduces neutrophil degranulation, whereas hyperinsulinemia inhibits fibrinolysis during human endotoxemia  [unique] | Investigated the impact of hyperglycemia and hyperinsulinemia in a human endotoxaemia model | 24  LPS (n = 24) | Healthy male volunteers | 22.8 ± 0.5  22.8 | 4 ng/kg  Escherichia coli  U. S. Pharmacopeial Convention  Rockville, MD | Temperature (oral)  Heart rate  Blood pressure  Oxygen saturation | TNF-α, IL-8, IL-10, and IL-6 | None | Cytokine levels increased  Neutrophils were activated  Induced endothelium and coagulation  Induced fibrinolysis inhibition | Blood samples were drawn up to 24 hours post-LPS | No safety concerns and dropouts indicated | The Netherlands |
| [49] Identification of candidate genes linking systemic inflammation to atherosclerosis; results of a human in vivo LPS infusion study  [unique] | Investigated genes connected to atherosclerosis in a human endotoxaemia model | 16  LPS (n = 11) | Healthy male volunteers | 23 ± 1  23 | 1 ng/kg  Escherichia coli  catalog number 1235503  lot G2B274  Pharmacopeial Convention, Inc,  Rockville | Temperature  The onset, duration, and clinical symptoms’ severity were monitored | IL-6 | None | Sickness symptoms were experienced by the volunteers (malaise, fever, chills, muscle ache, and nausea)  Monocyte counts reduced momentarily, and increased later  Differential expression was observed in the genes | Blood samples were taken up to 4 hours post-LPS | No safety concerns and dropouts indicated | The Netherlands |
| [50] Immunological and behavioral responses to in vivo lipopolysaccharide administration in young and healthy obese and normal-weight humans  [unique] | Investigated differences between healthy normal-weight and obese volunteers in a human endotoxaemia model | 40  LPS (n = 40)  Healthy obese (n = 14)  Healthy normal weight (n = 23) | Healthy obese and normal weight volunteers | 18 – 35  26.5 | Normal weight (0.8 ng/kg)  Obese (0.51 – 0.68 ng/kg)  Endotoxin Reference Standard  Escherichia coli  CAT number: 1235503  lot H0K354  United States Pharmacopeia  Rockville, MD | Blood pressure  Body temperature  Heart rate | IL-6, IL-10, and TNF-α | Sickness scores, mood state, state anxiety, and fatigue were assessed | Cytokine levels increased  Body temperature increased  Cortisol levels increased  Norepinephrine increased  Sickness symptoms increased  Fatigue increased  Negative mood increased  State anxiety increased | Up to 24 hours post-LPS | No safety concerns  3 exclusions reported, none due to LPS administration. | Germany |
| [51] Impact of training status on LPS-induced acute inflammation in humans  Trained (n = 8) - [unique] | Investigated how training state affected the ability of human volunteers to produce an inflammatory response triggered by LPS both systemically and in their adipose tissue and skeletal muscle. | 17  Untrained (n = 9) – [not unique]  Trained (n = 8) - [unique]  LPS (n = 8) | Healthy male volunteers | 23.8 ± 2.5  23.8 | 0.3 ng/kg  E. coli | Blood pressure  Body temperature  Heart rate | IL-6 and TNF-α | None | Heart rate increased  Leukocyte number increased  Neutrophil number increased  Monocyte number reduced  Cytokine levels increased  TNF- α mRNA levels in adipose tissue elevated  (reports are based on the effect of intravenous LPS in both trained and untrained volunteers) | Blood samples were collected up to 2 hours post-LPS | No safety concerns and dropouts indicated | Denmark |
| [52] Inflammation shapes neural processing of interoceptive fear predictors during extinction learning in healthy humans  [unique] | Investigated the core fear learning and extinction network's ability to develop and maintain conditioned fear in response to conditioned threats in a human endotoxaemia model | 95  LPS (n = 42)  Two studies were conducted.  46 volunteers – study 1  LPS (n = 21)  44 study - 2 LPS (n = 21) | Healthy volunteers | 18 – 45  31.5 | 0.4 ng/kg  Reference standard endotoxin  Escherichia coli  O113:H10  lot H0K354  United States Pharmacopeia  Rockville, MD | None mentioned | TNF-α and IL-6 | Assessed mood changes | Cytokine levels increased  Cortisol levels elevated  Positive mood decreased | Up to 24+ hours | No safety concerns  5 exclusions made (not reported as due to LPS injection) | Germany |
| [53] Inflammation-induced hyperalgesia: Effects of timing, dosage, and negative affect on somatic pain sensitivity in human experimental endotoxemia  [unique] | Investigated clinically relevant pain models, inflammatory indicators, pain sensitivity, and negative affect in relation to dose and time of LPS | 59  0.8 ng/kg LPS (n = 19)  0.4 ng/kg (n = 20)  LPS (LPS = 39) | Healthy male volunteers | 18 – 45  31.5 | 0.4 ng/kg and 0.8 ng/kg  Reference standard endotoxin  lot G3E069  United States Pharmacopeia  Rockville, MD | Blood pressure  Heart rate  Body temperature  (Used blood pressure cuff and aurical thermometer) | IL-6, IL-8, TNF-α, and IL-10 | State anxiety and mood were assessed | Cytokine levels increased (dose-dependent)  Body temperature increased (dose-dependent)  Cortisol levels increased  Positive mood self-reportedly declined  Calmness self-reportedly declined  Alertness self-reportedly declined  State anxiety increased  Pressure pain thresholds reduced (dose-dependent) | Blood samples were collected up to 6 hours post-LPS | No safety concerns  2 exclusions made (not reported as due to LPS injection) | Germany |
| [54] Inflammation-induced pain sensitization in men and women: does sex matter in experimental endotoxemia?  [unique] | Investigated the sensitization of pain in women and men after administering intravenous LPS | LPS (n = 40)  Men (n = 20)  Women (n = 20) | Healthy human volunteers  (Women volunteers were on hormonal contraceptives) | 18 – 45  31.5 | 0.4 ng/kg  Reference standard endotoxin  lot H0K354  United States Pharmacopeia  Rockville, MD | Heart rate (observed at the radial artery)  Body temperature (measured with a thermometer (intra-aurical))  Blood pressure (blood pressure cuff) | IL-6 and TNF-α | State anxiety was assessed  Side effect scores of LPS were rated | WBC counts increased  Pro-inflammatory cytokine levels elevated  Plasma cortisol levels elevated  Body temperature increased  Women exhibited a greater response in terms of cytokine, CRP and plasma cortisol levels. Nausea was reported as more pronounced among women than in men  State anxiety scores elevated | Blood samples were collected up to 6 hours post-LPS | No safety concerns and dropouts indicated | Germany |
| [55] Inhibition of protease-activated receptor (PAR1) reduces activation of the endothelium, coagulation, fibrinolysis and inflammation during human endotoxemia  [unique] | Investigated PAR1 inhibition on inflammatory response, coagulation activation, and activation of endothelia in a human endotoxaemia model | 16  LPS (n = 16) | Healthy volunteers | 18 and above  (reported as quartiles: 27 – 34 with a median age of 31)  30.5 | 2 ng/kg | None mentioned although vital parameters were reportedly measured | IL-6 and TNF-α | None | The inflammatory response was markedly decreased by PAR-1 inhibition | Blood samples were collected up to 24 hours post-LPS | One volunteer was excluded from the analysis phase (not reported as due to LPS)  LPS related adverse events were recorded (they were not reported as serious events). | Austria |
| [56] Insulin suppresses endotoxin-induced oxidative, nitrosative, and inflammatory stress in humans  [unique] | Investigated the effect of insulin on LPS-induced inflammatory responses  (these responses can be found in the key finding session) | 19  LPS (n = 9) | Healthy male volunteers | 20 – 33  26.5 | 2 ng/kg  Escherichia coli | Pulse  Blood pressure  Temperature | TNF-α, IL-6, and MCP-1 | Body aches, headaches, and chills scores were taken | Systolic pressure increased  Pulse rate increased  Temperature increased  Body aches, headaches, and chills scores elevated  Total leukocyte counts elevated  Nitric oxide metabolite levels increased  Levels of thiobarbituric acid–reacting substances elevated  The production of reactive oxygen species by polymorphonuclear leukocytes elevated  Free fatty acid levels in plasma elevated  Cytokine levels increased  CRP increased  Resistin and visfatin increased  Lipopolysaccharide binding protein levels increased  Macrophage migration inhibition factor elevated  High mobility group-B1 levels increased  Levels of myoglobin increased | Blood samples were collected up to 24 hours post-LPS | No safety concerns and dropouts indicated | USA |
| [57] Intravenous fish oil blunts the physiological response to endotoxin in healthy subjects  [unique] | Investigated intravenous fish oil’s effect on inflammatory responses in a human endotoxaemia model | 16  LPS (n = 16) | Healthy male volunteers | 18 – 35  26.5 | 2 ng/kg  USP  Rockville, Md  USA  lot no. 2 | Respiratory rate  Heart rate  Rectal temperature (Hellige, Servomed, from Germany)  Cardiac output (through thoracic bioimpedance, using NCCOM3 cardiodynamic monitor, BoMed, from Irvine California)  Arterial blood pressure (non-invasively measured using Critikon Dinamap from Tampa, Florida)  Transcutaneous oxygen saturation (from SpO2, measured with Pulse oximeter Biox 3740, from Ohmeda, Engelwood, California) | IL-6 and TNF-α | Nausea/vomiting, muscle ache, and headache | Volunteers reported experiencing headache, muscle pain, and nausea  Body temperature increased  Cardiac output increased  Endocrine response increased  ACTH levels increased  Cortisol levels increased  Glucagon increased  Plasma norepinephrine increased  Plasma epinephrine increased  IL-6 and TNF-α increased  Plasma lactate elevated  REE increased  Net fat oxidation elevated  FFA increased  (other effects can be found in the article) | Estimated at 6 hours post-LPS (360 minutes) | No safety concerns and dropouts indicated | Switzerland |
| [58] Intravenous Infusion of Human Adipose Mesenchymal Stem Cells Modifies the Host Response to Lipopolysaccharide in Humans: A Randomized, Single-Blind, Parallel Group, Placebo Controlled Trial  [unique] | Investigated the impact of allogenic adipose MSCs on inflammation in a human endotoxaemia model | 32  LPS (n = 32) | Healthy male volunteers | (could not report) | 2 ng/kg  Escherichia coli  U. S.  Standard reference endotoxin  National Institute of Health  Bethesda, ND | Blood pressure  Oral Temperature  Oxygen saturation (through the use of pulse oximetry)  Respiratory rate  Heart rate | TNF, IL-2, IL-4, IL-1β, IL-5, IL-6, IL-12p40, IL-8, IL-10, IL-13 | Sickness symptom scores were taken based on severity | Transient fever episode reported  Monocytopenia  Lymphocytopenia  Neutrophilic leukocytosis  Degranulation of neutrophil  IL-12p40, IL-6, IL-8, TNF and IL-10 levels increased  TATc levels increased  D-dimer increased  tPA levels increased  Soluble E-selectin increased  Soluble VCAM-1 increased  Soluble ICAM-1 levels increased  Plasma nucleosome concentrations increased  Gene expressions altered | Blood samples were drawn up to 24 hours post-LPS | No dropouts indicated  Six adverse events recorded (none reportedly serious) in five volunteers. | The Netherlands |
| [59] Pain perception in healthy volunteers: effect of repeated exposure to experimental systemic inflammation  [unique] | Investigated the effect of intravenous LPS administration on pain perception in healthy male volunteers | 17/20  LPS (n = 17) | Healthy male volunteers | 18 – 35  26.5 | 2 ng/kg  Lot EC-6  National Institutes of Health  Bethesda, MD | Heart rate  Pulse oximetry  Blood pressure (non-invasively determined)  Temperature | IL-1β, IL-8, IL-10, TNF- α, and IL-6 | Assessed pain | Sickness symptoms were experienced  Body temperature increased  TNF- α and IL-6 levels increased  Cortisol elevated  Pain pressure threshold decreased  Pain perception increased | Blood samples were collected up to 6 hours post-LPS | Three dropouts reported (none related to LPS)  No safety concerns | Denmark |
| [60] Lipopolysaccharide-induced experimental immune activation does not impair memory functions in humans  [unique] | Investigated the cognitive effects of LPS-injection into healthy male volunteers | 24  LPS (n = 12) | Healthy male volunteers | 18 – 38  28 | 0.4 ng/kg  Escherichia coli endotoxin  Serotype O113:H10  Lot G3E069  United States Pharmacopeia  Rockville, Maryland | Temperature (ear thermometer)  Heart rate  Blood pressure (blood pressure cuff) | IL-6, IL-10 and TNF- α | Neuropsychological tests were performed | Body temperature increased  Feeling of sickness were experienced by volunteers as reported, without the emergence of any particular symptoms  Neutrophil counts increased  Cytokine levels increased  Free cortisol concentrations in saliva elevated  Cortisol concentrations in plasma elevated  ACTH levels in plasma increased  Norepinephrine levels increased  Concluded, LPS had no effect on cognitive processing, memory, or attention | Blood samples were collected up to 6 hours post-LPS | No safety concerns and dropouts indicated | Germany |
| [61] Low dose LPS does not increase TLR4 expression on monocytes in a human in vivo model  [unique] | Investigated TLR4 and CD11b expression on human monocyte cell surfaces in a human endotoxaemia model | 16  LPS (n = 16) | Healthy male volunteers | (Could not report) | 0.4 ng/kg  E. coli LPS  0113:H10  United States Pharmacopeia | Body temperature  Heart rate  Blood pressure | IL-1β, IL-6, IL-10, IL-1ra, and TNFα | None | Circulating leukocyte concentrations increased  Marked lymphopenia  Lymphocyte levels decreased  Total number of granulocyte neutrophils elevated  Circulating monocytes decreased at first, then increased later  Cytokine levels increased  CD11b receptor density increased  Concluded, circulating mononuclear cells' surface expression of TLR4 remained unchanged after low-dose LPS injection. | Blood samples were collected up to 24 hours post-LPS | No safety concerns and dropouts indicated | Germany |
| [62] Low-dose steroid alters in vivo endotoxin-induced systemic inflammation but does not influence autonomic dysfunction  [unique] | Investigated the impact of a low-dose hydrocortisone infusion on the phenotypic characteristics, cytokine production, and HRV measures of endotoxin-induced systemic inflammatory responses. | 19  LPS (n = 19)  Male (n = 12)  Female (n = 7) | Healthy volunteers | 18 - 40  CORT+LPS (19 ± 0.3)  LPS (25 ± 2)  22 | 2 ng/kg  CC-RE  Lot #2 | Heart rate  Blood pressure  Core body temperature (rectal thermometer and oral) | IL-6, IL-8, IL-10, and TNF-α | None | Plasma cortisol concentrations increased  Temperature increased  Heart rate increased  IL-6, IL-10, and TNF-α levels increased  All assessed inflammatory mediators recovered to baseline levels 24 hours post-LPS  Respiratory rate increased  SDNN decreased  PNN_50_ decreased | Blood samples were drawn up to 24 hours post-LPS | No safety concerns and dropouts indicated | USA |
| [63] Mechanisms underlying the onset of oral lipid-induced skeletal muscle insulin resistance in humans  [unique] | Investigated the early phases of insulin resistance initiation caused by high-fat intake, compared to inducing a comparable level of insulin resistance in a human endotoxaemia model | 16  LPS (n = 16) | Healthy volunteers (lean insulin-sensitive) | 20 – 40  30 | 0.5 ng/kg  National Reference Endotoxin  Escherichia coli O:113  USP  Rockville, MD | Blood pressure  Body temperature  Heart rate | TNF-α, IL-6, and IL-1ra | None | Flu-like symptoms were experienced by the volunteers  Nonoxidative glucose use reduced  The proportion of granulocytes in blood leukocytes elevated  The percentage of lymphocytes reduced  Plasma cortisol levels increased  Cytokine levels increased  The associated genes’ mRNA expression elevated | Estimated at 8 hours on study day  (could not estimate using blood sampling) | No safety concerns and dropouts indicated | Germany |
| [64] Metabolic Alterations in Adipose Tissue During the Early Phase of Experimental Endotoxemia in Humans  [unique] | Investigated the metabolic activity of adipose tissue and its local blood flow in a human endotoxaemia model | 16  LPS (n = 8) | Healthy male volunteers | 22 – 38  30 | 4 ng/kg  Purified LPS  Escherichia coli O:113  Standard Reference Endotoxin  Food and Drug Administration  Bethesda, MD | Blood pressure (used plethysmography technique) measured with a Welch Ally medical device, Tycos Instruments, located at Arden, USA  Finger BP of the opposite arm (Finapres 2300, manufactured by Ohmeda, a company based in Louisville, USA)  Ventilation frequency via a respiratory-belt (ADinstruments GmbH, located in Germany)  Heart rate (3 lead ECG) | TNF-α, IL-6 and IL-1 | None | Flu-like symptoms were experienced by the volunteers  Body temperature increased  Mean blood pressure elevated  Blood flow increased proportionally to the adipose tissue located beneath the skin  Skin perfusion decreased  TNF-α and IL-6 levels increased  Cortisol levels increased  Adrenocorticotropin (ACTH) levels increased  Norepinephrine levels increased  Epinephrine levels increased  Catecholamine concentrations rose  Interstitial lactate elevated  Serum lactate elevated  Glycerol release from adipose tissue increased  Serum glycerol increased  Interstitial pyruvate increased  Serum pyruvate increased | (could not estimate from blood sampling)  Up to 3 hours post-LPS | No safety concerns and dropouts indicated | Germany |
| [65] MMP-8 Genotypes Influence the Inflammatory Response in Human Endotoxemia  [unique] | Investigated the impact of the rs1940475 single nucleotide polymorphism (SNP) on the downstream cytokine and chemokine response in healthy males challenged with LPS | 44  LPS (n = 44) | Healthy males (Caucasian) | Median age was 25.5 years, with an interquartile range (IQR) of 23 to 30 years  26.5 | 2 ng/kg  National Reference Endotoxin  Escherichia coli  CC-RE-Lot 2  National Institutes of Health  Bethesda, MD | ECG  Heart rate  Blood pressure  Oxygen saturation | MIP-1α, IL-6, IL-8 and TNF-α | None | Heart rate increased  MAP decreased  Body temperature increased  Cytokine levels increased  Plasma levels of thrombin-anti-thrombin complexes elevated  Prothrombin fragments F_1+2_ increased  Neutrophil counts increased  (other genotype specific results can be found in the article) | Blood samples were taken up to 24 hours post LPS | No safety concerns and dropouts indicated | Austria |
| [66] Neural Response to Emotional Stimuli During Experimental Human Endotoxemia  [unique] | Investigated the impact of LPS on the mood and brain responses of healthy male volunteers when exposed to emotionally upsetting visual stimuli.  Top of Form  Bottom of Form | 18  LPS (n = 18) | Healthy male volunteers | Age <18 or >40  29 | 0.4 ng/kg  Escherichia coli  United States Pharmacopeia  Lot G3E069 | Blood pressure  Temperature  Pulse | IL-6, IL-10, TNF-α, and IL-1ra | Mood parameters | Cytokine levels increased  Circulating neutrophil levels increased  Plasma cortisol levels elevated  Body temperature increased  Induced activation of the HPA-axis  Mood was impaired  Alertness declined  Calmness declined  State anxiety heightened | Blood samples were taken up to 24 hours post LPS | No safety concerns and dropouts indicated | Germany |
| [67] Nitric oxide inhalation and glucocorticoids as combined treatment in human experimental endotoxemia  [unique] | Investigated if administering low-dose iNO and glucocorticoids following LPS challenge would alter the inflammatory response in humans | 15  LPS (n = 15)  Women (n = 4)  Men (n = 11) | Healthy volunteers (white) | Average age (26.8 ± 1.4)  26.8 | 2 ng/kg  Endotoxin  Lot nr G3E0609  United States Pharmacopeia  Rockville, Maryland | Temperature (tympanic) – ThermoScan pro 1, device from Thermoscan, located at San Diego, California  Respiratory rate  SpO_2_  ECG  Blood pressure non-invasively measured (Datex Engström Light device, Helsinki, in Finland)  NO_2_ (INOvent, from Datex-Ohmeda, at Madison, Wisconsin) | TNF-α, IL-1ra, IL-6, IL-10 and IL-1β | Sickness symptom scores recorded | Flu-like symptoms were experienced  Volunteers reported tiredness and malaise  Body temperature increased  Heart rate increased  Diastolic pressure increased  WBC counts increased  Platelet counts reduced  TNF-a, IL-1ra, IL-6, and IL-10 levels increased | Blood samples were taken up to 5 hours | One dropout (a male volunteer) due to personal reasons (not reported as due to LPS)  No safety concerns | Sweden |
| [68] Effects of an Antisense Oligonucleotide Inhibitor of C-Reactive Protein  Synthesis on the Endotoxin Challenge Response in Healthy Human  Male Volunteers  [unique] | Assessed how the recent antisense C-reactive protein inhibitor, ISIS-CRP_Rx_, affects the inflammatory response during human endotoxaemia | 42  LPS (n = 30)  LPS (30/31) | Healthy male volunteers | 18 – 40  29 | 2 ng/kg  Escherichia coli O:113  United States Reference Standard Endotoxin  Clinical Center reference endotoxin  Lot 3 | ECG (12-lead)  Heart rate  Body temperature  Blood pressure | IL-1β, TNF-α, IL-6, and MCP-1 | Sickness symptoms were reported by volunteers | CRP levels increased  Pro-inflammatory cytokine levels increased (TNF-α, IL-6, and MCP-1)  WBC counts increased  Body temperature increased  Heart rate increased  Flu-like symptoms were experienced by all volunteers | Up to 72 hours post-LPS  (22 days for treatment and placebo) | Two volunteers dropped out before the endotoxin challenge  Adverse events recorded during follow-up (not reportedly due to LPS)  No safety concerns | USA |
| [69] Omega-3 PUFA supplementation and the response to evoked endotoxemia in healthy volunteers  [unique] | Investigated the impact of fish-oil supplement (Omega-3 PUFA) on inflammatory responses in a human endotoxaemia model | 60  LPS (n = 60) | Healthy volunteers | 18 – 45  31.5 | 0.6 ng/kg  Standard reference endotoxin  lot No. CCRE-LOT-1 +2  Clinical Center  Pharmacy Department  National Institutes of Health  Bethesda, MD | Heart rate  Temperature  Blood pressure | TNF-α, IL-6, MCP-1/CCL2, IL-10 and IL-1RA | None | Inflammatory response was experienced by volunteers  Body temperature increased  Cytokine levels increased  CRP levels increased | Blood samples were collected up to 24 hours post-LPS | No dropouts for volunteers that received LPS. Reported dropouts were before the LPS administration  No safety concerns | USA |
| [70] Pharmacokinetics and pharmacodynamics of the dual FII/FX inhibitor BIBT 986 in  endotoxin-induced coagulation  [unique] | Evaluated the impact of three doses of BIBT 986 in a human endotoxaemia model | 48  LPS (n = 48)  (analysis on 44 reported) | Healthy male volunteers | 18 – 40  29 | 2 ng/kg | Body temperature | TNF-α, and IL-6 | None | Flu-like symptoms were experienced and recorded as adverse events  F_1+2_ levels increased  APPT reduced  Collagen epinephrine reduced  Closure periods of adenosine diphosphate reduced  Platelet counts reduced  Cytokine levels increased  P-selectin activated  Activity in the fibrinolytic system elevated  Body temperature elevated  WBC counts change induced | Up to 48 hours | Four volunteers were excluded (at the end of the trial)  No safety concerns and dropouts indicated | Austria |
| [71] In vivo evidence for nitric oxide–mediated calcium-activated potassium-channel activation during human endotoxemia  [unique] | Investigated if the sensitivity to norepinephrine could be regained amidst human endotoxemia | 36  LPS (n = 36)  Women (n = 18), Men (n = 18)  [main study] | Non-smoking volunteers | (could not estimate) | 2 ng/kg  U. S. Standard Reference Endotoxin  E coli O:113  USP  Rockville, Md | Heart rate  Blood pressure  Body temperature measured using a tympanic thermometer (from Sherwood Medical, located at ‘s-Hertogenbosch, in the Netherlands) | Interferon-γ, TNF-α, IL-1β, IL-6, and IL-10 | None | Flu-like symptoms were experienced by the volunteers  Body temperature elevated  WBC counts reduced momentarily, then increased later  CRP levels increased  Cytokine levels increased  Blood pressure reduced  Heart rate elevated  FBF increased  Vasoconstriction induced by the administration of norepinephrine hindered | Up to 22 hours post-LPS | No safety concerns and dropouts indicated | The Netherlands |
| [72] Potent irreversible P2Y_12_ inhibition does not reduce LPS-induced coagulation activation in a randomized, double-blind, placebo-controlled trial  [unique] | Investigated the impacts of prasugrel intake in a human endotoxaemia model | 20  LPS (n = 16) | Healthy male volunteers | 18 – 40  29 | 2 ng/kg  National Reference Endotoxin  Escherichia coli  CC-RE-Lot 2  National Institutes of Health | (Did not indicate which specific vital signs were monitored)  *Temperature | None | None | Flu-like symptoms were experienced  Body temperature increased  Clotting time reduced (as well as CFT, and CT+CFT)  MCF increased  Histone-complexed DNA (hcDNA) elevated  F1+2 increased  TAT increased  P-selectin elevated  MPTF increased  Formation of monocyte-platelet aggregates diminished  Formation of the complete granulocyte-platelet aggregate increased | Blood samples were collected up to 24 hours post-LPS | No safety concerns and dropouts indicated | Austria |
| [73] Pretreatment with stress cortisol enhances the human systemic inflammatory response to bacterial endotoxin  [unique] | Studied the impact of stress cortisol pretreatment in a human endotoxaemia model | 36  LPS (n = 36) | Healthy volunteers | 18 – 55  36.5 | 2 ng/kg  Escherichia coli endotoxin  Clinical Center Reference Endotoxin  Lot 67801  Pharmacy Development Section  NIH  Bethesda, MD | ECG  Pulse oximetry  Blood pressure  Core temperature | TNF-α, IL-10, and IL-6 | None | Heart rate increased  Body temperature increased  Flu-like symptoms were experienced by the volunteers  Hypotension was absent in volunteers  Cortisol concentrations in plasma elevated  Adrenocorticotropic hormone levels in plasma increased  Plasma cytokine levels increased  C-reactive protein levels increased  Blood monocyte counts (total periphery) reduced | (could not estimate)  *Up to 8 hours | No safety concerns and dropouts indicated | USA |
| [74] Race and gender variation in response to evoked inflammation  [unique] | Compared and analyzed the variations in inflammatory responses by gender and race | 294  LPS (n = 294)  Main study | Healthy volunteers  (Male, female, African Ancestry and European Ancestry) | 18 – 45  31.5 | 1 ng/kg  E coli  U.S. standard reference  lot No. CCRE-LOT-1+2  Clinical Center  Pharmacy Department  National Institutes of Health  Bethesda, MD | Temperature  Blood pressure  Heart rate | TNF-α, IL-6, and IL-1RA | None | Heart rate increased  Temperature increased  Blood pressure (diastolic/systolic) increased*  Cytokine levels increased  CRP increased  Serum Amyloid A (SAA) increased  (Increase in cytokine was low in AA, same trend in SAA and CRP)  WBC counts increased  (CRP increased more in males) | Blood samples were drawn up to 24 hours post-LPS | No safety concerns and dropouts indicated | USA |
| [75] Reactivity of retinal blood flow to 100% oxygen breathing after lipopolysaccharide administration in healthy subjects  [unique] | Investigated the reactiveness of retinal vascular in a human endotoxaemia model | 18  LPS (n = 18) | Healthy male volunteers | 27.2 ± 3.8  27.2 | 20 IU/kg  National Reference Endotoxin  Escherichia coli  United States  Pharmacopeial Convention Inc.  Rockville, MD | Blood pressure (systolic, diastolic, and MAP)  Pulse rate  Oxygen saturation  (measured non-invasively via a Careview System from Hewlett Packard, located at Palo Alto, in California)  Temperature (Tympanic – Thermoscan device, from San Diego, California) | None | None | Sickness symptoms were experienced by the volunteers  Pulse rate increased  Leukocyte counts increased  Blood pressure decreased  Changed heart rate and body temperature (elevation)  WBC density increased  WBC flux increased  VD and AD elevated  RBC flux elevated | (could not estimate duration) ~ 4 hours LPS* | No safety concerns and dropouts indicated | Austria |
| [76] Reparixin, a specific interleukin-8 inhibitor, has no effects on inflammation during endotoxemia  [unique] | Investigated the effects of reparixin in a human endotoxaemia model | 20  LPS (n = 20) | Healthy male volunteers | 18 – 40  29 | 2 ng/kg  National reference endotoxin  Escherichia coli  USP Convention  Rockville, MD | Blood pressure  Oxygen saturation  Pulse rate | IL-6, TNF-α, and IL-8 | None | Body temperature increased  Heart rate reduced, then increased later  MAP reduced  Neutrophil counts elevated  Monocyte counts decreased  Lymphocyte counts declined  The cyclooxygenase pathway augmented  Thromboxane synthesis induced  F_1+2_ levels elevated  Cytokine levels elevated  The amount of CXCR1 and 2 (IL-8 receptors) on neutrophils in circulation reduced  Mean fluorescence intensity (MFI) of CD11b (degranulation marker) increased | Blood samples were drawn up to 24 hours | No safety concerns and dropouts indicated | Austria |
| [77] Response to systemic endotoxemia among humans bearing polymorphisms of the Toll-like receptor 4 (hTLR4)  [unique] | Compared hTLR4 genotypes in a human endotoxaemia model | 57  Male (n = 38)  Female (n = 19)  LPS (n = 57) | Healthy volunteers | 20 – 40  30 | 2 ng/kg  CCRE Lot 2 | Temperature  MAP  Heart rate | TNF-α and IL-6 | Symptom score* | Heart rate increased  Temperature increased  WBC counts increased  MAP decreased  (as seen from the figures in the article) | Up to +24 hours | No safety concerns and dropouts indicated | USA |
| [78] Reversal of immunoparalysis in humans in vivo: A double-blind, placebo-controlled, randomized pilot study  [unique]  [revised] | Investigated the immunoparalytic effects of GM-CSF and IFN-γ in a human endotoxaemia model  [main study] | 18  LPS (n =18) | Healthy male volunteers | 18 – 35  26.5 | 2 ng/kg  E. coli | Heart rate  Temperature  MAP | IL-10, IL-1RA, TNF-α and IL-6, IFN-γ, and granulocyte macrophage colony-stimulating factor (GM-CSF) | Symptom scores | Sickness symptoms (flu-like) were experienced by the volunteers  Cytokine levels increased  Momentarily decreased leukocyte counts, then later increased  Leucopenia  Heart rate increased  Temperature increased  MAP reduced | 8 – 24 hours post-LPS | No safety concerns and dropouts indicated | The Netherlands |
| [79] Selective increase of cerebrospinal fluid IL-6 during experimental systemic inflammation in humans: association with depressive symptoms  [unique] | Investigated the difference in cytokine responses in both CSF and plasma, and also assessed changes in mood in a human endotoxaemia model | 18  LPS (n = 10) | Healthy male volunteers | 20 – 42  31 | 0.8 ng/kg  Reference standard endotoxin  Escherichia coli  lot H0K354  United States Pharmacopeia  Rockville, MD | Heart rate  Arterial blood pressure  (measured with a Siemens Sc 900 XL device, from Dräger Medical, located at Lübeck, in Germany)  Body temperature (used a tympanic thermometer from a Genius 2 by Covidien, located at Tullamore, in Ireland | IL-6, TNF-α, IL-1β, and IL-10 | Mood assessed | Body temperature increased  CRP levels in serum increased  Leukocytosis induced  IL-6, IL-10, and TNF-α levels increased  Mood worsened  Cytokine response in CSF varied from the response in plasma | Blood samples were collected up to 24 hours post-LPS | No safety concerns and dropouts indicated | Germany |
| [80] Sickness behavior is not all about the immune response: Possible roles of expectations and prediction errors in the worry of being sick  [unique] | In a human endotoxaemia model, sickness expectations of volunteers were compared with LPS-induced sickness behavior and emotional state | 22  LPS (n = 22) | Healthy volunteers | 19 -34  26.5 | 2 ng/kg  Escherichia coli endotoxin  Lot H0K354  CAT number 1235503  United States Pharmacopeia  Rockville, MD | None mentioned | IL-6, IL-8, and TNF-α | Sickness and emotional behaviors were assessed | IL-6 levels increased  Sickness behavior increased  State anxiety increased  Emotional arousal reduced  Negative affect reduced | Blood samples were drawn up to 7 hours post-LPS | No safety concerns and dropouts indicated  *Data from a volunteer was omitted from analyses (not LPS related) | Sweden |
| [81] Systemic inflammation enhances stimulant-induced striatal dopamine elevation in tobacco smokers  [unique] | Investigated methylphenidate-induced elevation in dopamine in a human endotoxaemia model | Smokers (8)  Healthy control (n = 8) – Previous study  Total (used 16)  LPS (n = 8) | Tobacco smokers  Healthy volunteers | Tobacco smokers (32 ± 3.6)  32 | 0.8 ng/kg | Heart rate  Blood pressure  Temperature | TNF-α, IL-6, and IL-8 | Sickness scores were assessed | Cytokine levels increased (same way in both groups)  (other results were reported) | Blood samples were collected up to 3.5 hours post-LPS | No safety concerns and dropouts indicated  One exclusion made (not LPS-related) | USA |
| [82] Temporal metabolic profiling of plasma during endotoxemia in humans  [unique] | Investigated the patterns present in the metabolome of plasma in a human endotoxaemia model | 19  LPS (n = 15) – 11 men, 4 women | Healthy volunteers | 18 – 40  Mean age (22.7)  22.7 | 2 ng/kg  Clinical Center reference endotoxin  National Institutes of Health | None mentioned | None mentioned | None | Metabolic changes induced | Blood samples were collected up to 24 hours | No safety concerns and dropouts indicated | USA |
| [83] The Central Inflammatory Network: A Hypothalamic fMRI Study of Experimental Endotoxemia in Humans  [unique] | Established the hypothalamic regions responsible for detecting inflammation and examined how they connect with set brain networks on a broader scale | 7  LPS (n = 7) | Healthy normotensive males | 26.0 ± 8.5  26.0 | 1 ng/kg  GMP-grade LPS  Escherichia coli  O:113:H10:K-strain  Lot 94332B1  National Institute of Health  Clinical Center  Bethesda, MD | Body temperature (orally measured)  Pulse (by photoplethysmography)  Respiratory frequency and amplitude  Blood pressure (through digital artery) | TNF and IL-1β | Sickness and anxiety scores were assessed | TNF levels increased  ACTH levels increased  Leukocyte counts increased | Discharged volunteers 6-8 hours post-LPS*  Blood samples were collected up to 6 hours | One volunteer excluded (developed inflammation pre-LPS)  Another exhibited higher flu-like symptoms (temperature of 37.5°C) | Germany |
| [84] The Influence of Concentration/Meditation on Autonomic Nervous System Activity and the Innate Immune Response: A Case Study  [comparison group data from multiple studies]  Unique – single volunteer | Studied the influence of the meditation of a volunteer on ANS activity and immune response (innate). Also, compared these responses to others in another group (comparison group) | Single volunteer [unique}  Comparison group (n = 112)  LPS (n = 1) | Single volunteer (Dutch male)  Healthy male volunteers | Single volunteer (aged: 51)  51 | 2 ng/kg | Temperature | TNF-α, IL-10 and IL-6 | Symptom scores | Cytokine levels increased  Cortisol levels increased  HRV indices reduced | Up to 8 hours post-LPS | No safety concerns and dropouts indicated | The Netherlands |
| [85] The selective sirtuin 1 activator SRT2104 reduces endotoxin-induced cytokine release and coagulation activation in humans  [unique] | Investigated the effects of SRT2104 in a human endotoxaemia model | 24  LPS (n = 24) | Healthy nonsmoking male volunteers (Caucasian) | 22.9 ± 0.5  22.9 | 4 ng/kg  Escherichia coli  lot #118884  U.S. standard reference endotoxin  National Institutes of Health  Bethesda, MD | None mentioned | IL-6, IL-10, IL-8, IL-1β and TNF-α | None | Plasma levels of TNF-α, IL-10, and IL-6 increased  CXC chemokine IL-8 increased  Flu-like symptoms were induced  F1+2 levels increased  TATc increased  von Willebrand factor concentrations in plasma increased  tPA levels elevated  Plasminogen activator inhibitor-1 elevated  Elastase-α1-antitrypsin concentrations increased | Blood samples were drawn up to up to 21 hours post-LPS | No safety concerns and dropouts indicated | The Netherlands |
| [86] The effects of cold exposure training and a breathing exercise on the inflammatory response in humans: a pilot study  [Multiple studies, but; unique]  [Unique] | Studied, in a human endotoxaemia model, inflammatory response alterations due to breathing exercise and cold exposure | LPS study (n = 48)  Non-LPS (n = 40)  LPS (n = 48) | Healthy male volunteers | 21 (19 – 24)  *Median and interquartile range  21.5 | 2 ng/kg  Escherichia coli 0:113  Clinical Center Reference Endotoxin  Pharmaceutical Development Section  NIH  Bethesda, MD | Heart rate (by a 3-lead ECG)  Blood pressure (through an intra-arterial canula)  Respiratory rate & Oxygen saturation (by pulse oximetry) – (used a Philips MP50 patient monitoring device, from Eindhoven, located in the Netherlands)  Body temperature (recorded by a tympanic thermometer, First-Temp Genius device, from Sherwood Medical, located at Norfolk, Nebraska in the United States) | IP-10, IL-6, IL-10, IL-8, MIP -1 α, MCP-1, MIP-1β and TNF-α | Symptoms scores were assessed | Plasma epinephrine concentrations increased  Heart rate increased  Temperature increased  Symptoms scores increased  Cytokine levels increased | Up to 8 hours post-LPS | No safety concerns and dropouts indicated | The Netherlands |
| [87] Transcompartmental inflammatory responses in humans: iv versus endobronchial administration of endotoxin  [unique] | Compared intravenous LPS administration to endobronchial instillation in a human model of endotoxaemia | 15  LPS (n = 15) | Healthy male volunteers (nonsmoking) | 23 ± 2  23 | 4 ng/kg  Escherichia coli | Blood pressure (invasively measured)  Heart rate  Respiratory rate  Pulse oximetry (arterial)  Rectal Temperature | TNF-α and IL-6 | Sickness scores measured | Fever was induced  Blood pressure decreased  Tachycardia  Respiratory rate elevated  Systemic symptom scores elevated  Pulmonary symptom scores increased  Cytokine levels increased  Total peripheral leukocytes elevated  Neutrophils increased  CRP increased  BALF leukocyte counts increased  BALF IL-6 concentrations increased, similar for TNF-α | Blood samples were collected at 24 hours post-LPS | No safety concerns and dropouts indicated | Denmark |
| [88] Transcriptomic predictors of inflammation-induced  depressed mood  [unique]  (Treat it as unique since inclusion criteria did not capture any of the other published articles that used this data) | Studied whether baseline activity of transcription factors could predict depressive mood in a human endotoxaemia model | 115  LPS (n = 58) | Healthy volunteers | 18 – 50  Mean age (24.2)  24.2 | 0.8 ng/kg  Escherichia coli  Group 0:113 | None mentioned | (Could not determine) | Sickness symptom scores and depressed mood were assessed | Self-reported depressed mood elevated | ~ 6 hours post-LPS | No safety concerns and dropouts indicated  (case of one missing volunteer data) | USA |
| [89] Transfusion of 35-day-stored red blood cells does not alter lipopolysaccharide tolerance during human endotoxemia  [main published article is not part of the included articles]  [uniquely captured study]  [unique] | Studied the impact of transfused RBCs (stored for 35 days) on cytokine response in a human endotoxaemia model | 18  LPS (n = 18) | Healthy volunteers | 18 – 35  26.5 | 2 ng/kg  National Institute of Health  Clinical Center | *Blood pressure  *Heart rate (reported tachycardia)  *Respiratory rate (Tachypnea)  *Possibly measured | Transforming growth factor (TGF)-β, IL-6, IL-8, IL-1β, and TNF-α | None | Blood pressure reduced  Tachycardia  Tachypnea  Fever  Leukocytosis  Cytokine levels increased  Biphasic pattern induced (typical to LPS) | Completed the study 8 hours post-LPS | No safety concerns and dropouts indicated | The Netherlands |
| [90] Transvenous vagus nerve stimulation does not modulate the innate immune response during experimental human endotoxemia: a randomized controlled study  [unique] | Investigated tVNS in a human model of endotoxaemia | 20  LPS (n = 20) | (Non-smoking) healthy males | Sham group (24 (22 - 26))  tVNS group (26 (23 - 27))  25 | 2 ng/kg  Purified LPS  U. S. Standard Reference Endotoxin  Escherichia coli 0:113  Pharmaceutical Development Section  National Institutes of Health  Bethesda, MD | Arterial blood pressure  Heart rate (3-lead ECG)  Temperature measured by an infrared tympanic thermometer (a First-Temp Genius device, from Sherwood Medical, at Crawley/Sussex, in the United Kingdom) | TNF-α, IL-10, IL-6, and IL-8 | Symptoms scores were assessed | Body temperature increased  Heart rate increased  Flu-like symptoms induced  MAP reduced  Cytokines levels increased  Leukocytosis | ~ 8 hours post-LPS | No safety concerns and dropouts indicated | The Netherlands |
| [91] Treatment with acetylsalicylic acid reverses endotoxin tolerance in humans in vivo: a randomized placebo-controlled study  [unique] | Investigated the impact of acetylsalicylic acid (ASA) on tolerance induced by endotoxin in a human model of endotoxaemia | Healthy males (n = 30)  Sepsis patients (n = 4)  LPS (n = 30) | Healthy males  Sepsis patients | 18 – 35  26.5 | Initial bolus of 1 ng/kg, then 1 ng/kg/hr. | *Temperature  *Blood pressure  *Heart rate | IL-6, IL-10, MIP-1 α, IL-8, IL-1RA, TNF-α, MCP-1, and MIP-1β  IL-1β, IL-13, and IL-4 | Symptoms scores were assessed | Cytokine levels increased (first endotoxin dose)  Dampened cytokine release (second dose – endotoxin tolerance)  Urinary PGE-M levels elevated  Body temperature increased  Symptom scores increased  Heart rate elevated  MAP decreased | ~ 8 hours post-LPS | No safety concerns and dropouts indicated | The Netherlands |
| [92] Tumor necrosis factor-α inhibition protects against endotoxin-induced endothelial glycocalyx perturbation  [unique] | Studied the influence of LPS-induced inflammation on the thickness of endothelial glycocalyx | 21  LPS (n = 21) | Healthy male volunteers (Caucasian) | Etanercept (24 ± 4)  Saline (23 ± 3)  23.5 | 1 ng/kg  Escherichia coli LPS  Catalog number 1235503  Lot G2B274  U.S Pharmacopeial Convention Inc.,  Rockville, MD | Blood pressure  Body temperature  Heart rate | IL-6 | None | Clinical symptoms were experienced by the volunteers  Blood pressure reduced  Heart rate increased  Body temperature elevated  Glycocalyx thickness in the microvasculature was decreased  Density of the capillary reduced  Hyaluronan concentrations in plasma increased  Hyaluronidase activity in plasma reduced  CRP and IL-6 levels increased  Monocyte counts decreased  The portion of monocytes expressing CD62L^+^ reduced  F1+2 increased  D-dimer increased | ~ 24 hours post-LPS | No safety concerns and dropouts indicated | The Netherlands |
| [93] Type 2 diabetes mellitus is associated with impaired cytokine response and adhesion molecule expression in human endotoxemia  [unique] | Investigated, in a human endotoxaemia model, the difference between diabetic and non-diabetic inflammatory responses | 42  Healthy (n = 23)  Type 2 Diabetes Mellitus (n = 19)  LPS (n = 42) | Male volunteers (Healthy and Type 2 Diabetes Mellitus) | Healthy – 59 (53 - 65)  Type 2 Diabetes Mellitus – 55 (48 - 62)  * Mean (95% CI)  57 | 0.3 ng/kg  E. coli LPS  Lot G2 B274  United States Pharmacopeial Convention  Rockville, MD | Heart rate  Blood pressure  Temperature (Tympanic) | IL-1ra, IL-6, and TNF | None | WBC count induced  Neutrophil levels increased  Lymphocyte counts declined  Monocytes declined initially then increased  Temperature increased  Heart rate elevated (highly elevated among diabetic volunteers)  Cytokine levels increased  E-selectin elevated  VCAM-1 increased  ICAM-1 increased | ~ 8 hours post-LPS | No safety concerns and dropouts indicated | Denmark |
| [94] Upregulation of renal inducible nitric oxide synthase during human endotoxemia and sepsis is associated with proximal tubule injury  [unique] | Studied, in a human endotoxaemia model, the elevation of iNOS | Septic patients (n = 10)  Controls (n = 11)  (8 + 14)  LPS (n = 22) | Septic patients and healthy volunteers | Septic patient (56.5 ± 5.9)  Controls (22.1 ± 0.7)  LPS (Protocol 1) – (23.9 ± 1.0)  LPS (Protocol 2) – (23.0 ± 0.7)  LPS and aminoguanidine – (22.3 ± 0.7)  23.07 | 2 ng/kg  Escherichia coli LPS  E. coli O:113  United States Pharmacopeia Convention  Rockville, MD  *8 volunteers (Protocol 1) | Heart rate  Blood pressure  Body temperature | TNF-α and IL-10 | None | Flu-like symptoms were experienced by the volunteers  Body temperature increased  CRP elevated  Cytokine levels increased  Mean heart rate elevated  MAP reduced  (other LPS-induced effects were reported in the article) | ~ 24 hours | No safety concerns and dropouts indicated  *Raised concerns but from another article. | The Netherlands |
| [95] VPAC1 receptor expression in peripheral blood  mononuclear cells in a human endotoxemia model  [unique] | Investigated the receptor expression of VPAC1 in PBMC during human endotoxaemia | 20  LPS (n = 20) | Healthy male volunteers | 18 – 45  31.5 | 2 ng/kg  E. coli endotoxin  U.S. Standard Reference Endotoxin  NIH Clinical Center  Bethesda, MD | Mean blood pressure  Body temperature  Heart rate | None mentioned | None | Flu-like symptoms experienced by the volunteers  Body temperature elevated  BP reduced  Heart rate elevated  WBC counts elevated  Neutrophil counts elevated  Monocyte counts reduced  Lymphocyte cell counts decreased  VPAC-1 positive cells reduced  Receptor expression reduced initially then elevated later  Plasma levels of VIP elevated | Blood samples were drawn up to 24 hours | No safety concerns and dropouts indicated | Austria |
| [96] High-density lipoprotein attenuates inflammation and coagulation response on endotoxin challenge in humans  [unique] | Investigated, in a human endotoxemia model, the relation between low high -density lipoprotein and increased sensitivity to LPS, an inflammatory stimulus | 27  (13 + 14)  LPS (n = 27) | Healthy male volunteers | Low HDL (35.4 ± 2.5)  Normal/High HDL (35.1 ± 5.2)  35.25 | 1 ng/kg  Escherichia coli lipopolysaccharide  catalog number  1235503, lot G2B274  United States Pharmacopeial Convention Inc,  Rockville, Md | Body temperature  Heart rate  Blood pressure | TNF-α, IL-1β, MCP-1, IL-6, and IL-8 | Symptoms scores were recorded | Flu-like symptoms experienced by the volunteers  Heart rate increased  Low-density lipoprotein cholesterol elevated  Monocytes and leukocytes increased  Neutrophil response increased  Serum cytokine levels increased  CRP levels increased  LPS binding protein concentrations elevated  Prothrombin fragments (F_1+2_) increased  Fibronolytic markers elevated | Blood samples were drawn up to 24 hours | No safety concerns and dropouts indicated | The Netherlands |
| [97] Endotoxin tolerance does not limit mild ischemia-reperfusion injury in humans in vivo  [unique] | Investigated the effect of pretreating with LPS, in incremental order, against injury due to ischemia-perfusion | 24  LPS (n = 14) | Normotensive, non-smoking males | 23 ± 4  23 | Incremental dosages (0.2, 0.5, 1.0, 2.0 ng/kg)  Reference Escherichia coli LPS  (lot Ec-5;  Center for Biologic Evaluation and Research, FDA  Bethesda, MD, USA | Temperature using a tympanic thermometer | TNF-a, IL-6 and IL-10 | Symptoms scores were recorded | Five consecutive days of endotoxin administration induced endotoxin tolerance in humans  Dose dependent alteration in temperature and symptom score levels  WBC counts increased  Plasma cytokines increased dose-dependently | Blood samples were drawn up till 6 hours post LPS | One exclusion was made due to suspected viral infection  No safety concerns mentioned | The Netherlands |
| [98] A human model of inflammatory cardio-metabolic dysfunction; a double blind placebo-controlled crossover trial  [unique] | Utilized low dose LPS (0.6 ng/kg) to induce minimal changes in metabolic and inflammatory responses | 10  LPS (n = 10) | Non-smoking volunteers (male and female) | 22.7 ± 3.8  22.7 | 0.6 ng/kg  Standard reference endotoxin.  lot# CC-RE-LOT-  1 + 2  Clinical Center, National Institutes of Health  US. | Body temperature  Heart rate  Blood pressure | TNF-a, and IL-6 | Assessed pain | Plasma cytokines increased  CRP levels increased  LPS-placebo comparison showed no substantial changes in temperature  WBC counts increased  Heart rate increased  Serum cortisol and growth hormone increased slightly  mRNA levels of multiple inflammatory genes elevated  Induced systemic insulin resistance | ~ 24 hours | No safety concerns mentioned | USA |
| [99] A randomized study of the efficacy and safety of intravenous acetaminophen compared to oral acetaminophen for the treatment of fever  [unique] | Investigated the effects of two routes (IV and oral) of acetaminophen administration on LPS-induced fever | Enrolled 105 volunteers    81 completed the study  LPS (n = 105) | Healthy males | 33.0 ± 10.52  33.0 | Test IV dose – 1 ng/kg  Study dose - 4 ng/kg  Reference Standard Endotoxin  E. coli 0:113  NIH  U.S | Core temperature was measured using a “Jonah Ingestible Core Temperature Capsule” [99]. | None | None | Mean core temperature increased in the two groups (IV and PO)  Fever was induced  Liver function tests (LFTs) were slightly elevated | ~ 6 hours | No safety concerns  24 withdrawals were made due to vomiting  Two volunteers in the IV acetaminophen experienced nausea and diarrhea – maybe related to the administered study drug  Aspartate aminotransferase (AST) and Alanine aminotransferase (ALT) percentages elevated | USA |
| [100] Amino acid supplementation is anabolic during the acute phase of endotoxin-induced inflammation: A human randomized crossover trial  [unique] | Investigated the influence of amino acid supplementation on LPS-induced catabolism | 8  LPS (n = 8) | Healthy lean males | 25 – 32  28.5 | 1 ng/kg  E. coli  10,000 USP endotoxin  Lot HOK354  US Pharmacopeial Convention Inc.  Rockville, MD | Axillar temperature  ECG  Non-invasive blood pressure  Heart rate | TNF-a, IL-1 β, IL-10 and IL-6 | None | Volunteers experienced flu-like symptoms (headache, discomfort, chills, and palpitations)  Stress hormones and growth hormones increased (Cortisol, glucagon)  Serum cytokine levels increased  Temperature increased  Heart rate increased  Whole body protein synthesis increased | Blood samples were collected up till the 6^th^ hour post-LPS | No safety concerns mentioned | Denmark |
| [101] Bupropion pre-treatment of endotoxin-induced depressive symptoms  [unique] | Investigated the influence of pre-treating volunteers with bupropion on LPS-induced symptoms | 10  LPS (n = 10)  Male (n = 5)  Female (n = 5) | Healthy volunteers | - | 0.8 ng/kg | Heart rate  Heart rhythm  Body temperature  Blood pressure | Il-8, IP-1, MCP-1, AND MPI-1β | Assessed depression, social interest, fatigue and vigor | Heart rate increased  Body temperature increased  Serum biomarker levels (cytokines and chemokines) increased  Depressive symptoms increased  Fatigue increased  Vigor decreased  Social interest decreased | Blood samples were collected up till the 4^th^ hour post-LPS | 28 volunteers were screened, 8 were deemed ineligible and 7 dropped out after screening. 13 began the study but three failed to return after the first study condition.  One and 9 volunteers completed two and three study conditions respectively.  Out of the 10 final participants, one fell ill and did not complete one of the conditions  No safety concerns mentioned | USA |
| [102] C-reactive protein is expressed and secreted by peripheral blood  mononuclear cells  [unique] | Investigated CRP in peripheral blood mononuclear cells (PBMC) expression through IV LPS in human.volunteers | 6  LPS (n = 6) | Healthy male volunteers | 19 – 28  23.5 | 2 ng/kg  National reference endotoxin  Escherichia coli  USP  U.S Pharmacopeia Convention Inc.,  Rockville,  MD, USA | Heart rate  Body temperature  Blood pressure | *In vitro (IL-1a, TNF-a,  IL-6 and IL-10) | None | Volunteers experienced flu-like symptoms  Pulse rate increased  Body temperature increased  CRP levels increased  Neutrophil levels increased  Monocyte an lymphocyte levels reduced | Volunteers were discharged between 8 to 10 hours post-LPS | No safety concerns mentioned | Austria |
| [103] Cerebral net exchange of large neutral amino  acids after lipopolysaccharide infusion in healthy  humans  [unique] | Investigated the influence of IV LPS infusion on the plasma ratio  between branched-chain and aromatic amino acids (BCAA/AAA ratio) | 12  LPS (n = 12) | Healthy male volunteers | 20 – 33  26.5 | 0.075 ng/kg/h  (total 0.3 ng/kg) – 4 hr* 0.075 ng/kg  Batch G2 B274  US Pharmacopeial Convention  Rockville, MD  USA | Heart rate  Blood pressure  Capillary oxygen saturation | TNF-α | None | Arterial PCO_2_ reduced  pH increased  CMR of oxygen CMRO_2_ slightly elevated  Intermittent dozing among some volunteers  Plasma phenylalanine increased | Volunteers were released after 12 hours | No safety concerns mentioned | Denmark |
| [104] Changes in HLA-DR expression, cytokine production  and coagulation following endotoxin infusion in  healthy human volunteers  [unique] | Investigated, in a human endotoxemia model, the effects of IV LPS on HLA-DR expression, coagulation activation, and cytokine production | 9  LPS (n = 9) | Healthy male volunteers | - | 3 ng/kg  USP standard endotoxin  Escherichia coli | Mean sitting blood pressure  Heart rate  Oral temperature  ECG (continuous lead II)  12-lead ECG  Pulse oximetry | IL-6, IL-1ra, IL-2, IL-10, IL-8, G-CSF, GM-CSF, IL-1β, sTNF receptor I, TNF-α | None | Serum cytokine levels increased (TNF-α, sTNFR, IL-6, IL-8, IL-1Ra, IL-1β, IL-10)  WBC counts increased  Volunteers experienced flu-like symptoms (lethargy, pyrexia, myalgia, back pain, rigors, headache, dizziness, and paraesthesia)  Mean sitting blood pressure elevated  Oral temperature increased  Heart rate increased  Coagulation and fibrinolysis were induced  Liver serum enzymes increased  Aspartate  aminotransferase (AST) increased  alanine aminotransferase (ALT) increased  GGT (gamma glutamyl transpeptidase increased  CRP increased  total bilirubin, lactate dehydrogenase (LDH) and  glucose increased from baseline | ~ 48 hours | No safety concerns mentioned | UK |
| [105] Circulating and muscle glutathione turnover in human endotoxaemia  [unique] | Investigated the kinetics of glutathione in a human endotoxemia model | 8  LPS (n = 8) | Healthy male volunteers | 23 ± 3  23 | 4 ng/kg  U.S Standard Reference  E. coli endotoxin  Lot EC-6  US Pharmacopeia | Heart rate  ECG  Blood pressure  Oxygen saturation  (Datex-Engstrom light monitor)  Temperature (in the outer ear) | - | None | Sickness symptoms were exhibited by the volunteers (headache, shivering, muscle pain, nausea in some volunteers and malaise)  Low-grade fever occurred  Total glutathione reduced | ~ 8 hours | No safety concerns mentioned | Sweden |
| [106] Citalopram reduces endotoxin-induced fatigue  [unique] | Pre-treated volunteers with citalopram to investigate its effect on LPS-induced depressive symptoms | 11  LPS (n = 11) | Healthy volunteers  Female (n = 5)  Male (n = 6) | 32 ± 9  32 | 0.8 ng/kg  Reference endotoxin | Heart rate  Heart rhythm  Blood pressure  Body temperature | TNF and IL-6 | Assessed behavior (depression, anxiety) | Heart rate increased  Serum biomarker levels increased  Interest in social interactions reduced | Blood samples were collected up till the 3^rd^ hour post-LPS | One volunteer experienced nausea and withdrew participation  No safety concerns mentioned | USA |
| [107] Comparison of host immune responses to LPS in human using an immune profiling panel, in vivo endotoxemia versus ex vivo stimulation  [unique] | Used an immune profiling panel (IPP) to compare immune responses to LPS both in vivo and ex vivo | 8  LPS (n = 8)  (There was ex vivo experiment with whole blood from 8 healthy volunteers) | Non-smoking healthy male | 18 – 25  21.5 | 2 ng/kg  E. coli 0113 reference Endotoxin  NIH  Bethesda, MD  USA | None | IL-10, IL-1β, IL1RN, IP10, IL-18, IL-2, IFN-γ, TNF-α | None | Monocyte, lymphocyte, and eosinophil counts decreased  Neutrophil counts increased  Cytokines (TNF-α, IL-1β, and IFN-γ) were under-expressed during ex vivo LPS compared to in vivo | ~ 4 hours post-LPS | No safety concerns mentioned | Netherlands |
| [108] Development of endotoxin tolerance in humans *in vivo*  [unique] | Investigated, in a human endotoxemia model, the development of endotoxin tolerance after repeated E. coli LPS administrations for 5 days | 14  LPS (n = 14) | Healthy male volunteers | 22 ± 2  22 | 2 ng/kg/day for 5  (1 minute)  *Escherichia coli* LPS  US Reference  Lot EC-5 endotoxin  CBER, FDA  Bethesda, MD | Tympanic temperature  Heart rate (ECG)  Blood pressure/MAP | Proinflammatory cytokines (TNF-α, IL-6, IL-1β)  Anti-inflammatory cytokines (IL-10, IL-1RA, and TGF- β) | Symptoms scores were collected (headache, nausea, backache, shivering, and muscle ache) | Volunteers experienced flu-like symptoms  Symptom scores increased  Blood pressure decreased  Heart rate and temperature increased  In subsequent LPS administrations, symptom score, temperature, blood pressure, and heart rate were substantially less pronounced  Creatinine concentrations slightly elevated, then significantly during repeated LPS administrations which peaked on day 3 and returned to baseline on day 5  WBC counts initially decreased, then elevated by the 6th hour. It was attenuated on day 5  C-reactive proteins peaked on day 3 and returned to baseline values by day 5  Monocytes demonstrated higher values on day 5, 4 hours post-LPS | 5-day study | No safety concerns mentioned | The Netherlands |
| [109] Diffusion-weighted MR spectroscopy (DW-MRS) is sensitive to LPS-induced changes in human glial morphometry: A preliminary study  [unique] | In a human endotoxemia study, researchers investigated the apparent diffusion coefficients (ADC) of metabolites of glia and neuron | 7  LPS (n = 7) | Healthy male volunteers | 25.3 ± 5.9  25.3 | 1 ng/kg  Escherichia coli O:113  US Standard Reference Endotoxin  NIH | Heart rate  Body temperature | TNF-α, IL-6, and IL-10 | Self-rated mood and sickness scores | Heart rate increased  Body temperature increased  Cytokine levels increased  Substantial momentarily changes in mood, sickness symptoms, and fatigue  Fatigue and sickness score increased  WBC counts increased  Glial metabolite choline’s ADC substantially elevated | 2 weeks study duration  Blood samples were collected 6 hours post-LPS | No safety concerns mentioned | UK |
| [110] Direct effects of locally administered lipopolysaccharide on glucose, lipid, and protein metabolism in the placebo-controlled, bilaterally infused human leg  [unique] | In an infused leg LPS model, researchers investigated insulin resistance and protein/lipid metabolism | 8 | Healthy male volunteers | 27.5 ± 1.0  27.5 | 0.025 ng/kg/h (continuously administered over 360 minutes (6 hours)  USP Endotoxin  Lot G3E069  US Pharmacopeia Convention, Inc.  Rockville, MD | Mean blood pressure  Heart rate | TNF-α, GM-CSF, IL-2, IL-5, IL-4, IL-6, IL-10, IL-8, and INF- γ, IL-1β | None | Palmitate release increased  Insulin-stimulated glucose uptake was inhibited | > 6 hours | No safety concerns mentioned | Denmark |
| [111] The LPS-Induced Increase in Circulating Microparticles is Not Affected by Vitamin C in Humans  [unique] | Investigated the influence of administered high-dose vitamin C on the formation of microparticles in a human endotoxemia model | 14  LPS (n = 14) | Healthy, non-smoking, and drug-free | 19 – 40  Median = 26  29.5 | 2 ng/kg  E. coli  US Pharmacopeia Convention Inc.  Rockville, MD | Body temperature | None | None | Volunteers experienced sickness symptoms (headache, fever, muscle pain, vertigo, arthralgia, and systemic hypotension)  Circulating levels of vitamin C decreased  D-dimer increased  CRP increased  Body temperature increased  Leucocyte counts increased | Blood samples were collected 6 hours post-LPS | No safety concerns mentioned | Austria |
| [112] Protein Kinetics in Human Endotoxaemia and Their Temporal Relation to Metabolic, Endocrine, and Proinflammatory Cytokine Responses  [unique] | Investigated the kinetics of whole-body protein in a human endotoxemia model | 6  LPS (n = 6) | Healthy males | 27 – 39  Mean age – 32  32 | 4 ng/kg  Lot EC-6  Escherichia coli 0113  USPC  Rockville, Maryland  USA | Temperature  Heart rate  Mean arterial pressure  Energy expenditure  Glucose oxidation rate  Fat oxidation rate | TNFα and IL-6, IL-1β | None | The volunteers experienced fever and tachycardia  Resting energy expenditure elevated  Lipid oxidation rates increased  Plasma glucose rose  Plasma glycerol increased  Non-esterified fatty acid levels increased  Plasma cortisol levels increased  TNFα and IL-6 levels increased  Plasma growth hormone increased | Arterialized venous blood samples were collected up till 600 minutes (10 hours) | No safety concerns mentioned | UK |
| [113] Nicotine Exposure Alters In Vivo Human Responses to Endotoxin  [unique] | Studied the effects of nicotine on LPS-induced responses | 12  LPS (n = 11) | Adult normal males | 18 – 40  29 | 2 ng/kg  Clinical Center Reference Endotoxin  CC-RE-Lot 2  NIH | Rectal temperature  Mean arterial pressure (MAP)  Heart rate | TNF-a, IL-6 and IL-8 | Assessed sickness symptoms (nausea, muscle aches, headache, chills, perceived sensitivity to light and fever) | Volunteers experienced sickness symptoms  Body temperature increased  WBC count initially dropped, subsequently increased  Plasma cortisol levels increased | Discharged volunteers on day 3 (24-hour post LPS) | One volunteer was dropped for suspected tobacco use  No safety concerns mentioned | USA |
| [114] Effects of a Cytokine Inhibitor, JTE-607, on the Response to Endotoxin in Healthy Human Volunteers  [unique] | Investigated JTE-607’s cytokine attenuation effects in a human endotoxemia model | 27  LPS (n = 9) | Healthy Caucasian males | 18–45  31.5 | 3 ng/kg  E. coli | 12-lead ECG  Continuous lead II (ECG)  Pulse oximetry | sTNF receptor, IL-6, IL-8, IL-2,IL-1-ra, IL-10, G-CSF, GM-CSF, TNF-α, IL-1 β | None | IL-1ra, IL-10, IL-6, TNF-α, CRP, IL-8 levels increased  IL-1β increased modestly | > 24 hours post-LPS | Adverse events included, vomiting dizziness, pyrexia, rigors, back pain, myalgia, lethargy, paresthesia, feeling hot, headache, and nausea  No safety concerns mentioned | UK |
| [115] β-Lactoglobulin is insulinotropic compared with casein and whey protein ingestion during catabolic conditions in men in a double-blinded randomized crossover trial.  [unique] | Compare muscle protein kinetics and metabolism after ingestion of β-lactoglobulin, casein, and whey. Used LPS to mimic/cause inflammatory state | 9  [LPS = 9] | Healthy males, BMI 20-30, no regular medications | 20-40  30 | Bolus of LPS (1 ng/kg) from E. coli (10,000 USP endotoxin, lot HOK354; US Pharmacopeia Convention) | Blood pressure, heart rate, axillary temperature | TNF-α, IL-6, CRP | Shivering, headache, nausea, muscle pain | TNF-α and IL-6 peaked two hours after LPS infusion and came almost back to baseline after 6 hours. CRP was elevated 24 hours after injection. Temperature rose about two degrees C 4 hours after injection, then almost leveled out the following day. Heart rate rose about 25 bpm at 4 hours, then almost leveled out the next day. MAP briefly peaked after 2 hours, then decreased to below baseline after 3 hours. Leveled out by 24 hours post injection. | Hourly until 6 hours post-LPS, then 24 hours after injection | None | Denmark |
| [116] Tissue Factor Expression in Monocyte Subsets During Human Immunothrombosis, Endotoxemia, and Sepsis  [unique] | Examining tissue factor expression by monocytes and its role in sepsis-induced coagulopathy | 13  LPS (n = 13) | Healthy volunteers | 18-37  27.5 | Bolus of LPS (2ng/kg); Clinical Center Reference Endotoxin facility (Lot 94332B1) | None mentioned | CRP | None mentioned | CRP increased | 7 days | None | UK |
| [117] The Effect of Glutamine Infusion on the Inflammatory Response and HSP70 During Human Experimental Endotoxaemia  [unique] | Examine the effect of glutamine on the cytokine response | 8  LPS (n = 8) | Healthy men, unremarkable medical history, no regular medications, normal pre-screening | 21-33  27 | IV bolus of E. coli endotoxin (0.3ng/kg); Lot EC-6, Pharmacopeia Convention, Rockville, MD, USA | Rectal temperature (continuous), heart rate (continuous), EKG (continuous), BP (q15min), venous oxygen saturation | TNF-α, IL-6 | None mentioned | TNF-α (peak 4 hours) and IL-6 (peak 5 hours) increased after endotoxin, both leveled back out by hour 10; temperature increased (peak at hour 5 after LPS), heart rate increased (peak at hour 4 after LPS) | Hourly for 10 hours (2 w/o LPS, 8 w/ LPS) on 2 separate trial days (one placebo, one intervention) separated by a 30-day washout; blood draws hourly and at 0.5 and 1.5 hours after LPS | None | Denmark |
| [118] Systemic Administration of Oxytocin Reduces Basal and Lipopolysaccharide-Induced Ghrelin Levels in Healthy Men  [unique] | Examining the role of oxytocin administration on ghrelin concentration during endotoxemia. | 10  LPS (n = 10) | Healthy men, normal fasting glucose and lipids, liver, kidney, thyroid, and heme function | 20-40  30 | 2 ng/kg, National Reference, E. coli endotoxin, USP Convention, Rockville, MD, USA | None mentioned | None mentioned | None mentioned | None | 6.5 hours | None | Austria |
| [119] Skeletal Muscle Contractile Properties and Proinflammatory Cytokine Gene Expression in Human Endotoxaemia  [Unique] | Examines the mechanism for muscle weakness during endotoxemia. | 12  LPS (n = 12) | Healthy men, no history or sign of cardiorespiratory, metaboli, or inflammatory disease, no medication or injury for 3 months before trial | 18-45  31.5 | E. coli LPS 4ng/kg IV over 10 minutes, Lot EC-6, USPC, Rockville, Maryland, USA | Pulse ox (continuous), EKG (continuously), brachial BP (q30min), rectal temp (continuous) | TNF-α, IL-6 | Prodromal malaise 60-90 minutes after LPS; somnolence, photophobia, headaches, rigors after 90-180 minutes | HR rose (peak around 3-4 hours post-LPS), temp rose (peak around 3-4 hours), MAP rose (peak around 2 hours); TNF-α rose (peaked around 2 hours, then decreased), IL-6 rose (peaked around 3 hours, then decreased) | 6 hours | None | UK |
| [120] Plasma Fractalkine is a Sustained Marker of Disease Severity and Outcome in Sepsis Patients  [unique]  [cannot state with certainty] | Establish whether plasma fractalkine levels are elevated in sepsis and associated with outcomes | 5 | Healthy, non-smoking males | 19-22  20.5 | 4 ng/kg LPS from E. coli O:113, CC-RE lot 3, NIH, Bethesda, MD | None mentioned | IL-6 mentioned briefly? | None mentioned | None mentioned | 20 hours | None | The Netherlands |
| [121] Muscle Mitochondrial Activity Increases Rapidly After an Endotoxin Challenge in Human Volunteers  [unique] | Using endotoxemia to study muscle mitochondrial function in early sepsis | 7  LPS (n = 7) | Healthy males, good health | 23-29  26 | IV endotoxin, E. coli, Lot EC-6, US Pharmacopenia, Rockville, MD, 4 ng/kg | Heart rate (continuous), EKG (continuous), O2 sat (continuous), BP (continuous), temperature (hourly) | None mentioned | Shivering, headache, muscle pain, malaise, nausea | Temperature rose (peak around 3 hours), O2 sat remained stable, HR rose (peak at 4 hours), MAP rose briefly after 2 hours, then dropped at 4 by 4 hours | 4 hours after LPS | None | Sweden |
| [122] Metabolic and Physiologic Effects of and Endotoxin Challenge in Healthy Obese Subjects  [unique] | To test physiologic response of healthy obese subjects to LPS | 8  LPS (n = 8) | BMI 30.5-38.9, without glucose intolerance and lipid abnormalities, no medication, <10 cigarettes per day  (Male = 4, and female = 4)  Healthy obese | 27-41  34 | 2 ng/kg IV bolus endotoxin from E. coli, batch no. 2 USP, Rockville, MD | Rectal temperature, heart rate, respiratory rate, arterial BP, cardiac output, oxygen saturation | TNF-α, IL-6, hs-CRP | Myalgia, headache, shivering, malaise, nausea, vomiting | Temperature rose (peak around 2.5 hours, then back to baseline), TNF-α rose (peak at 2 hours, then back near baseline), IL-6 rose (peaked around 2 hours, not back to baseline by 4 hours), hs-CRP rose slightly (peak at 6 hours), heart rate increased, BP decreased but not significant, cardiac output increased | Two different days, 8 hours each, with 2 weeks separation | None | Switzerland |
| [123] Melatonin Suppresses Markers of Inflammation and Oxidative Damage in a Human Daytime Endotoxemia Model  [unique] | Examine effects of melatonin on inflammation | 12  LPS (n = 12) | Healthy males, non-smoking, no AUD, no medications, no melatonin allergy, no known sleep difficulties, and no injections in last 14 days | 18-40  29 | 0.3 ng/kg IV bolus E. coli LPS endotoxin (Lot G3E069; US Pharmacopeia Convention, Rockville, MD) | None mentioned | TNF-α, IL-1β, IL-6, YKL-40, IL-1Ra, IL-10 |  | TNF-α peaked for both groups at 2 hours, IL-1β stayed about the same until it rose in hour 8 for placebo, IL-6 peaked at 2 hours for both groups, YKL-40 steadily rose for both groups over 8 hours, IL-1Ra peaked at 4 hours for both groups, IL-10 peaked at 2 hours for both groups | 8 hours; two days separated by 3 weeks | None | Denmark |
| [124] Macrophage Migration Inhibitory Factor (MIF) in Meningococcal Septic Shock and Experimental Human Endotoxemia  [unique] | Studied MIF and cytokines in experimental endotoxemia | 8  LPS (n = 8) | Healthy | 18-25  21.5 | 2 ng/kg IV E. coli O:113 LPS (batch O:113, United States Pharmacopia Convention Rockville, MD) | Temperature, heart rate, blood pressure | TNF-α, IL-1β, IL-10 | Nausea | Pyrexia, increased heart rate, drop in BP, TNF-α peaked after 1.5 hours after LPS, IL-1β and IL-10 increased slightly after 2-3 hours | 22 hours after injection | None | The Netherlands |
| [125] Low-Dose Endotoxin Potentiates Capsaicin-Induced Pain in Man: Evidence for a Pain Neuroimmune Connection  [unique] | Searching for evidence that immune activation has a role in development and maintenance of chronic pain in humans | 12  LPS (n = 12) | Good general health, men, non-smokers | 20-41  30.5 | 0.4 ng/kg IV; Reference Standard endotoxin Lot G3E069, EC-6, E. coli O113:H10:K(-), US Pharmacopeia, Rockville, MD, USA | Tympanic temperature (q15m for 1 hour, then hourly for 5 more hours), core temperature (continuous) | IL-1β, IL-6, IL-10, TNF-α | Headache, nausea, vomiting, chills, lethargy | Increased temperature and heart rate; increase in IL-6 with peak at 3 hours; no significant change in IL-1β, TNF-α, or IL-10 | 6 hours | None | Australia |
| [126] Low Plasma Gelsolin Concentrations in Chronic Granulomatous Disease    [unique]  (Age was not mentioned) | Exploring concentration of plasma gelsolin in those with CGD. Comparing with inflammation caused by endotoxemia. | 54 (unclear if all healthy volunteers got endotoxin, but seems like it)  LPS (n = 4) | Some had CGD, some healthy controls  Healthy | Not mentioned | 4 ng/kg IV GMP endotoxin prepared from E. coli O113 by List Biological Laboratories Inc., Campbell CA | Temperature | TNF-α | None mentioned | Temperature peaked at 2.5 hours and returned to baseline by 24 hours; TNF-α peaked at 1-1.5 hours | 24 hours | None | USA |
| [127] Lipopolysaccharide Infusion Enhances Dynamic Cerebral Autoregulation Without Affecting Cerebral Oxygen Vasoreactivity in Health Volunteers  [unique] | Assess the impact of isocapnic hypoxia and hyperoxia on dynamic cerebral autoregulation during early stages of sepsis | 10  LPS (n = 10) | Healthy, non-sedentary, male. Unremarkable medical history, no signs of infection in last 4 weeks, no regular medication | 21-25 (SD, not range)  23 | 4 hour LPS infusion, 0.5 ng/kg/hour; E. coli LPS batch G2 B274, US Pharmacopeial Convention, Rockville, MD, USA) | Heart rate (EKG), invasive BP, O2 sat, respiratory rate, rectal temperature (baseline, then hourly), pH | TNF-α | Malaise, shivering, headache, myalgia, dizziness, nausea | TNF-α peaked 1 hour after cessation of infusion; IL-6 also increased; increase in heart rate, MAP unaffected; in normoxia group, PaO2 decreased after LPS, PaCO2 also decreased, pH increased | 6 hours | None | Denmark |
| [128] Left Ventricular Diastolic Filling Characteristics Are Not Impaired But Systolic Performance Was Augmented in the Early Hours of Experimental Endotoxemia in Humans  [unique] | To determine whether endotoxemia causes diastolic dysfunction. | 11  LPS (n = 11) | Normotensive, healthy  (7 men and 4 women) | 24-36  30 | IV E. coli endotoxin (4 ng/kg); standard reference endotoxin, Bureau of Biologics, Food and Drug Administration, Bethesda, MD | EKG (continuous), BP, pulse ox, temperature (hourly) | None mentioned | None mentioned | Temperature, HR, cardiac index increased significantly; systolic, diastolic, and MAP decreased significantly | 5 hours | None | USA |
| [129] Intravenous Administration of LPS Activates the Kynurenine Pathway in Healthy Male Human Subjects: A Prospective Placebo-Controlled Cross-Over Trial  [unique] | Investigate how LPS administration affects the kynurenine pathway. | 10  LPS (n = 10) | Non-smoking men free from medication, without known history of relevant disease | 20.4-27.8  24.1 | IV LPS over 5 min. (E. coli O113, 2 ng/kg); National Reference Bacterial Endotoxin, lot #94332B1, Investigational Drug Management at the National Institutes of Health (NIH), Bethesda, MD) | EKG, heart rate, non-invasice BP, temperature | IL-6 | None mentioned | Increased temperature and heart rate, decreased systolic and diastolic BP | 6 hours | None | Austria |
| [130] Influence of Tedizolid on the Cytokine Response to the Endotoxin Challenge in Healthy Volunteers: A Cross-Over Trial  [unique] | Investigate the influence of tedizolid on the cytokine response to endotoxin challenge. | 15  LPS (n = 14) | Healthy males, normal history and physical, non-smoking | Not mentioned  Inclusion allowed (18 – 55)  36.5 | LPS, E. coli. O113, 2 ng/kg over 1-2 min.; Reference Endotoxin, CC-RE Lot 3; NIH, Bethesda, MD, USA | Body temperature, blood pressure, heart rate, O2 sat, ECG | IL-6, TNF-α | Headache, flu-like symptoms, vertigo | IL-6 and TNF-α peaked after 2 hours in both groups; mentions tachycardia and hypotension as adverse events, but no data given | 6 week washout, 24 hours | 1 dropped out because of compliance issues and was replaced | Austria |
| [131] In Vivo Lipopolysaccharide Exposure of Human Blood Leukocytes Induces Cross-Tolerance to Multiple TLR Ligands  [unique] | Investigate whether exposure of humans to LPS induces tolerance in circulating leukocytes to other TLR agonists | 6  LPS (n = 6) | Healthy males | 25-35  30 | Bolus IV LPS 4 ng/kg (E. coli, lot G, US Pharmacopeia) | None mentioned | TNF, IL-1β, IL-6, IL-10 | None mentioned | Transient release of TNF (peak at 90 and returned to baseline after 4 hours), IL-6 (peak at 4 hours), and IL-10 (peak at 3 hours), but not IL-1β | 24 hours | None | The Netherlands |
| [132] In Vivo Endotoxin Synchronizes and Suppresses Clock Gene Expression in Human Peripheral Blood Leukocytes  [unique] | Determine the state of clock gene expression in leukocytes during endotoxin challenge. | 4  LPS (n = 4) | Male and female | 18-29  23.5 | 2 ng/kg endotoxin (NIH Clinical Center Reference endotoxin, CC-RE, Lot 2, Bethesda, MD) | None mentioned | TNF-α, IL-6 | None mentioned | TNF-α peaked withinh 1-1.5 hours postinfusion and returned to baseline by 3 hours, IL-6 peaked within 2 hours and returned within 6 | 24 hours | None | USA |
| [133] Immunomodulatory effects of fosfomycin in experimental human endotoxemia  [unique] | Explore the immunomodulatory effects of Fosfomycin in a human endotoxemia study | 12  LPS (n = 12) | Healthy males | 18 – 40  29 | 2 ng/kg | None mentioned | IL-6, IL-1, and TNF-a | None mentioned | Volunteers experienced sickness symptoms (chills, fever, and headache) | 24 hours | None | Austria |
| [134] Human Models of Low-Grade Inflammation: Bolus versus Continuous Infusion of Endotoxin  [unique] | To create a model of systemic low-grade inflammation with bolus or continuous infusion of LPS. | 10  LPS (n = 10) | Healthy males. No medications, not infection in last 4 weeks. | 19-29 (SD)  24 | 0.3 ng/kg IV bolus E. coli endotoxin (batch G2 B274; US Pharmacopeial Convention, Inc. rockville, MD); 4 hour IV infusion of endotoxin (total dose 0.3 ng/kg) | MAP, heart rate, rectal temperature | TNF-α, IL-6 | None mentioned | Heart rate increased slightly after bolus (peak at 4 hours), but overall change in endotoxin groups was nonsignificant compared to placebo; overall slight increase in rectal temperature too small to be significant; no effect on MAP; TNF-α increased compared to placebo and increased more in the bolus than infusion group (peaks at 2 and 5 hours, respectively); IL-6 signifcantly increased only in bolus compared to placebo (peak at 3 hours and 6 hours, respectively) | 8 hours after LPS; three separate days | None | Denmark |
| [135] Human In Vivo Neuroimaging to Detect Reprogramming of the Cerebral Immune Response Following Repeated Systemic Inflammation  [unique] | Examined the immune response systemically and in the brain after repeated LPS challenge. | 6 (5)  LPS (n = 5) | Health males. Non-smokers, no medication | 18-35  26.5 | 2 ng/kg LPS IV bolus (E. coli type O113, lot no. 94332B1, List Biological Laboratories, Campbell, USA) | Heart rate (EKG), arterial BP, temperature | TNF-α, IL-6, IL-8, IL-10  IL-1RA, MCP-1, MIP-1a, MIP-1b* | Flu-like symptoms (headache, chills) | Temperature increased (peak around 3 hours), but more in first LPS dose; heart rate increased (peak around 4 hours), but more in first LPS dose; TNF-α peaked at 2 hours in first LPS dose, but only barely increased after second dose; IL-6 and IL-8 similar to TNF-α; IL-10 peaked around 3 hours, but more in first dose | 8 hours | One subject had baseline PET results that deviated more than 2 SD from the mean, so was considered an outlier and was excluded from analysis | The Netherlands |
| [136] Hematological Indices, Inflammatory Markers and Neutrophil CD64 Expression: Comparative Trends During Experimental Human Endotoxemia  [unique] | To examine expression of CD64 on neutrophils during inflammation. | 10  LPS (n = 10) | Healthy, non-smoking, no medications (except OCPs)  4 males  6 females | 18-24  21 | 2 ng/kg IV E. coli LPS (O113, US Pharmacopia, Rockville, MD, USA) | None mentioned | TNF-α, IL-6, IL-10, CRP, IFN-γ | None mentioned | CRP increased (peak at 22h); IFN-γ increased (peak at 1.5h); TNF-α increased (peak at 1.5h); IL-10 increased (peak at 2h); IL-6 increased (peak at 4h) | 22 hours | None | The Netherlands |
| [137] Genomic Responses in Mouse Models Poorly Mimic Human Inflammatory Diseases  [unique]  (not enough information to determine the uniqueness) | To demonstrate that mouse models of inflammation aren't similar to human responses. | 8  LPS (n = 4) | Healthy males and females  Female 1, males 3  Female 1, males 3 | 18-40  29 | 2 ng/kg IV NIH Clinical Center Reference Endotoxin E. coli O113 (CC-RE-lot 2) | None mentioned | None mentioned | None mentioned | None mentioned | 24 hours | None | USA |
| [138] Gene Expression Profiles of Peripheral Blood Leukocytes After Endotoxin Challenge in Humans  [unique] | To define gene expression in leukocytes after endotoxemia. | 8  LPS (n = 8) | Healthy males and females, no tobacco use | 20-45  32.5 | 4 ng/kg E. coli O:113 IV endotoxin (Clinical Center Reference Endotoxin, NIH, Bethesda, MD) | Temperature, heart rate, BP | TNF-α, IL-1β, IL-4, IL-5, IL-6, IL-8, IL-10, IL-12, IFN-γ | Malaise, headache, fever | Increased HR, decrease in MAP; TNF-α, IL-1β, IL-6, IL-8, and IL-10 rose at 3 hours and returned to baseline by 6-24 hours; IL-4 and IL-5 were not significant | 24 hours | None | USA |
| [139] Endotoxin-Induced Effects on Platelets and Monocytes in an in Vivo Model of Inflammation  [unique] | To investigate the effects of inflammation on platelet and monocyte activation. | 13  LPS (n = 13) | Healthy males, clinically asymptomatic, no cardiovascular disease of diabetes | 24-43  33.5 | IV LPS (2 ng/kg); National reference endotoxin E. coli; US Pharmacopeial Convention) | Body temperature, heart rate, BP | None mentioned | None mentioned | Temperature, heart rate, and BP remained unchanged after 24 hours; no details on in between values | 24 hours | None | Germany |
| [140] Endotoxemia Reduces Cerebral Perfusion but Enhances Dynamic Cerebrovascular Autoregulation at Reduced Arterial Carbon Dioxide Tension  [unique] | Examine the effects of endotoxin on dynamic cerebral autoregulation. | 10  LPS (n = 10) | Healthy, young. No medications or medical conditions | 24-40 (SD)  32 | Bolus IV E. coli endotoxin (2 ng/kg); US Pharmacopeia Convention, Rockville, MD | Temperature, MAP, heart rate, CO, PaCO2 | TNF-α | None mentioned | Temperature and TNF-α increased; PaCO2 decreased; no significant effect on MAP; increased cardiac output and heart rate | 3 hours | None | Denmark |
| [141] Effects of N-Acetylcysteine Against Systemic and Renal Hemodynamic Effects of Endotoxin in Healthy Humans  [unique] | Examine the influence of N-acetylcystein on cytokines, renal plasma flow, and systemic pressor response during endotoxemia. | 11 (8)  LPS (n = 8) | Healthy males, non-smokers, drug-free, no clinically relevant illnesses | 21-30  25.5 | IV E. coli LPS (2 ng/kg); National Reference Endotoxin E. coli USP Rockville, MD) | Temperature, heart rate, BP | TNF-α; IL-1β | None mentioned | MAP decreased on placebo days, but not NAC from 3-5 hours; in same interval, pulse increased for both groups, but went back to baseline after 7 hours for placebo but not NAC; increase in temperature for both groups between 5.5 and 7 hours, but returned to baseline only in NAC; TNF-α increased in both groups after 1.5 hours, but returned to baseline after 4 hours only in NAC; IL-1β increased 4 hours in both groups | 8 hours | 2 had adverse drug reactions to NAC and 1 took a different medication with salicylic acid | Austria |
| [142] Effects of Endotoxin on Lactate Metabolism in Humans  [unique] | To assess lactate production and clearance during LPS challenge. | 14  LPS (n = 14) | Health males, BMI between 19.5 and 25, no medications, no smoking | 18-30  24 | IV bolus LPS 2 ng/kg (US Pharmacopeial Convention, Rockville, MD) | Heart rate, rectal temperature, arterial pressure, cardiac output, O2 sat | TNF-α, IL-6, CRP | Headache, myalgia, nausea | Increase in temperature (peak at 4 hours) and heart rate; no change in blood pressure; TNF-α increased (peak at 90m); IL-6 increased (peak at 2-2.5h) | 6.5 hours | None | Switzerland |
| [143] Effects of Endotoxaemia on Markers of Permeability, Metabolism, and Inflammation in the Large Bowel of Healthy Subjects  [unique] | To assess the effects of endotoxemia on markers of permeability, metabolism, and inflammation in the large bowel. | 12  LPS (n = 12) | Healthy males with no medications or vaccines for 1 month. Non-smoker | 23.1-27.7 (SD)  25.4 | IV endotoxin (2 ng/kg); E. coli Lot EC-6, US Pharmacopeia Convention, Rockville, MD) | Temperature, heart rate (EKG), O2 sat, non-invasive BP | TNF-α, CRP | Nausea, shivering, malaise | Increase in temperature (peak at 3 hours) and heart rate (peak at 4 hours); no change in BP; TNF-α increased after 1 hour and normalized at 24 hours; CRP increased after 6 hours and did not normalize at 12 hours | 8 hours | None | Denmark |
| [144] Effect of Short-Term Intralipid Infusion on the Immune Response During Low-Dose Endotoxemia in Humans  [unique] | Explore the effect of short-term high levels of fatty acids on the inflammatory response. | 14  LPS (n = 14) | Healthy males, no history of medical problems | 24.2-26 (SD)  25.1 | Iv bolus of endotoxin (0.1 ng/kg); E. coli Lot G2 B274, US Pharmacopoeia Convention, Rockville, MD | Heart rate (EKG), noninvasive BP, temperature | IL-6, TNF-α | None mentioned | Temperature increased (no difference between groups); heart rate increased (no difference between groups); TNF-α increased in both groups (peak at 8 hours); IL-6 increased in both, but more in Intralipid (peak at 8 hours) | 11 hours | None | Denmark |
| [145] Effect of Melatonin on Human Nighttime Endotoxaemia: Randomized, Double-Blinded, Cross-Over Study  [unique] | To examine the anti-inflammatory effects of melatonin in nighttime endotoxemia. | 12  LPS (n = 12) | Healthy males. No smoking, alcohol abuse, medication, infections in last 14 days. | 19-31  25 | IV 0.3 ng/kg E. coli LPS; Lot G3E069, US Pharmacopeia Convention, Rockville, MD, USA | Blood pressure, temperature, heart rate | TNF-α, IL-1β, IL-1, IL-6, YKL-40, IL-1-RA, IL-10) | None mentioned | Increased levels of IL-1β, TNF-α, IL-6, IL-1Ra, IL-10, YKL-40, but not different between groups | 8 hours | None | Denmark |
| [146] Disassociation of Static and Dynamic Cerebral Autoregulatory Performance in Healthy Volunteers After Lipopolysaccharide Infusion and in Patients with Sepsis  [unique] | Assess cerebral autoregulation in a model of sepsis. | 9  LPS (n = 9) | Healthy males | 21-25 (SD)  23 | 4 hours infusion of E. coli LPS (batch G2 B274 US Pharmacopeial Convention, Rockville, MD)  0.5 ng/kg/h | Heart rate, invasive BP, O2 sat, rectal temperature | TNF-α, IL-6 | Flu-like symptoms | TNF-α peaked around hour 3 of infusion; IL-6 peaked at hour 4 of infusion; temperature peaked just after infusion; increased heart rate, temporary decrease in MAP; hyperventilation with decrease in PaCO2 and increased pH, no effect on CaO2 | 12 hours | None | Denmark |
| [147] Acute Experimental Inflammation in Healthy Women Attenuates Empathy for Psychological Pain  [unique] | To study endotoxemia specifically in women. | 52 | Healthy, all female. BMI 18-30, not menstruating during study | 18-40, mean for LPS group was 22.85 ± 3.18 | 0.4 ng/kg, reference standard endotoxin from E. coli O113:H10, lot H0K354, US Pharmacopeia, Rockville, MD | Blood pressure, heart rate, temperature, cortisol | TNF-α, IL-6, IL-10 | Mood, social interaction empathy task, sickness symptoms (GASE questionnaire) | Rise in temperature 3 and 4 hours post-injection, heart rate increased 2-6 hours post-injection. Increase in TNF-α, IL-6, IL-10, and cortisol. Sickness score increased 2-3 hours after LPS injection. Decrease in activity, positive mood. Increase in depressivity and fatigue. | 24 hours | None | Germany, Sweden |
| [148] Exposure to Normobaric Hypoxia Shapes the Acute Inflammatory Response in Human Whole Blood Cells in Vivo  [unique] | To study changes in gene expression in response to endotoxin challenge at hypoxic conditions. unique | 30 | Healthy males | 25.8 ± 2.9 | IV E coli HOK364, 0.4 ng/kg | Temperature, SpO2 | IL-6, TNF-α | Headache, mild shivering | Mild fever 3 hours post-injection, headache, mild shivering. Increased IL-6 and TNF-α | 24 hours | None | Germany, Sweden |
| [149] Neo-Epitope Detection Identifies Extracellular Matrix Turnover in Systemic Inflammation and Sepsis: An Exploratory Study  [unique] | To study ECM turnover in endotoxemia and sepsis. unique | 10 | Healthy males | 21 (19-22 range) | 2 ng/kg IV, US Standard Reference Endotoxin E. Coli O:113, NIH Bethesda, MD. |  | TNF-α, IL-6, IL-8, and IL-10 | None mentioned | None relevant mentioned | 24 hours | None | The Netherlands, Denmark |
| [150] Sick and Detached: Does Experimental Inflammation Impact on Movement Synchrony in Humans?  [unique] | Explore the effects of illness on movement synchrony. unique | 26 | Healthy female | 22.85 ± 3.18 | 0.4 ng/kg, | None mentioned | None mentioned | Mood, physical sickness symptoms | None relevant mentioned | 24 hours | None | Germany, Sweden |
| [151] Atazanavir-Induced Unconjugated Hyperbilirubinemia Prevents Vascular Hyporeactivity During Experimental Human Endotoxemia  [unique] | Investigate the impact of hyperbilirubinemia on antioxidant capacity, inflammation, and vascular dysfunction. unique | 20 | Healthy males, non-smokers | 22.35 | 2 ng/kg E. coli LPS IV | MAP, heart rate, temperature | TNF-α, IL-6, IL-8, IL-10, CRP | None mentioned | Cytokines all increased after LPS. Decrease in MAP, increase in heart rate, increase in temperature | 5 days | Started with 30, 5 had bilirubin raise too fast, two went vasovagal during cannula placement, 1 was unable to plan the experiment, 1 made an error with medication, and 1 had new 1st degree AV block BEFORE the LPS. Nothing related to LPS | The Netherlands |
| [152] Using a Wearable Patch to Develop a Digital Monitoring Biomarker of Inflammation in Response to LPS Challenge  [unique] | To determine a digital biomarker of inflammation using a wearable after endotoxemia. unique | 10 | Healthy (6 female and 4 male), non-smokers | 35.4 (22-45 range) | 1 ng/kg LPS IV | Pulse rate, respiratory rate, tympanic temperature | IL-6, IL-8, MCP-1, MIP-1α, MIP-1β, TNF-α | None mentioned | HR, RR, skin temp increased, HRV decreased. Cytokine levels relate to vitals to different extents, but no data on pure change in cytokines | 26 hours | None | USA, Belgium, The Netherlands, UK |
| [153] The Oral IRAK4 Inhibitors Zabedosertib and BAY1830839 Suppress Local and Systemic Immune Responses in a Randomized Trial in Healthy Male Volunteers  [unique] | Evaluate and characterize the pharmacological activity of IRAK4 inhibitors. unique | 51 | Healthy males | 19-55 | 1 ng/kg E. coli O113 LPS as a 2-min infusion | Pulse, BP, temperature | TNF-α, IL-6, IL-8, CRP, procalcitonin | Fever, chills, nausea, asthenia, spontaneous hematoma, myalgia, headache, oropharyngeal pain | TNF-α, IL-6, IL-8, CRP, and procalcitonin increased. Increased systolic BP, HR, and temperature. Fever, chills, nausea, asthenia, spontaneous hematoma, myalgia, headache, oropharyngeal pain. | 8 days | Three adverse effects of moderate intensity (tachycardia, infusion-related reactions of fever and chills) in two participants. LPS was discontinued. There was also one COVID and one URI that led to withdrawal. | Germany, The Netherlands |
| [154] Acute Hypoxic Conditions Preceding Endotoxin Administration Result in an Increased Proinflammatory Cytokine Response in Healthy Men  [unique] | To assess the impact of hypoxia on inflammation in men. unique | 36 | Healthy, male, Caucasian | 25.9 ± 2.9 | 0.4 ng/kg LPS IV | O2 sat, temperature, BP, heart rate, cortisol | IL-6, IL-8, TNF-α | None mentioned | LPS didn't affect O2 levels, temperature increased, no affect on BP or HR. Cortisol increased. IL-6, IL-8, and TNF-α all increased. | 8 hours | None | Germany, Sweden |
| [155] Non-Invasive Ventral Cervical Magnetoneurography as a Proxy of In Vivo Lipopolysaccharide-Induced Inflammation  [unique] | To measure compound action potentials in nerves during endotoxemia. unique | 11 | Healthy males | 20 (18-30 range) | 3 ng/kg LPS IV | Heart rate, HRV | TNF-α, IL-6, IL-8, IL-1beta, IL-10 | Likert scale for headache, nausea, rigor, and myalgia | Heart rate and HRV increased. All cytokines increased. Nausea, vomiting. | 7 hours | One subject passed out during IV insertion and was excluded. Two got nausea and vomiting so were excluded from analysis, but still got LPS. | USA |
